# Chronic Neurological and Psychiatric Outcome in TBI Patients Assessed by A classification System Integrating Symptoms and Radiological Evidence

**DOI:** 10.1101/2025.08.26.25334454

**Authors:** Ying Liu, Jie-Yu Wang, Huai-Yu Zhuchen, Jiale Chen, Yang Yu, Yi Wang, Zhi-Biao Huang, Cheng-Jia Yang, Guang-Quan Guo, Xiaojing Ye, Hu Zhao, Yan-Wei Shi, Yanni Zeng

**Affiliations:** Faculty of Forensic Medicine, Zhongshan School of Medicine, Sun Yat-sen University, Guangzhou, China; Guangdong Province Translational Forensic Medicine Engineering Technology Research Center, Zhongshan School of Medicine, Sun Yat-sen University, Guangzhou, China; Guangdong Province Key Laboratory of Brain Function and Disease, Zhongshan School of Medicine, Sun Yat-sen University, Guangzhou, China; Shenzhen Kangning Hospital, 1080 Cuizhu Rd, Luohu District, Shenzhen, China; Guangdong Mental Health Center, Guangdong Provincial People’s Hospital (Guangdong Academy of Medical Sciences), Southern Medical University, Guangzhou, China

## Abstract

**Background:** Traumatic brain injury (TBI) can lead to long-term cognitive and psychiatric consequences such as intellectual disability, depression, and anxiety. The mental outcomes result from a complex interplay between neurobiological, structural, and clinical factors that are unlikely to be fully reflected by a single measure focused on only one aspect. Profiling mental outcomes is essential for identifying TBI patients at risk of cognitive-psychiatric complications, ideally through a tailored assessment system that integrates both subjective and objective indicators, yet such a system is currently absent in clinical settings. Mental Disability Classification (MDC) is a TBI-specific measurement system for mental outcome, widely practiced by forensic psychiatrists in China, that incorporates both symptomatic and radiological indicators of mental impairment. Despite its advantage in integrating multiple types of evidence, its clinical application remains limited due to reliance on expert judgment and the absence of comprehensive evaluation. We aimed to establish and improve the MDC’s utility in clinical research by constructing prediction models, and profile the mental prognosis landscape of TBI patients through systematically evaluating the MDC system.

**Methods:** We performed a multicenter and longitudinal study of 499 TBI patients from four cohorts in China. SYSU-TBI1 cohort consisted of 356 TBI patients recruited from 2017 to 2019, from whom we collected 189 features including both acute and chronic phase variables concerning neurological and psychiatric symptoms, radiological characteristics, as well as demographic variables. SYSU-TBI1 cohort was used to construct prediction models for MDC using random forest and Least Absolute Shrinkage and Selection Operator (LASSO) methods, through which MDC’s key chronic dimensional features were also identified. SYSU-TBI1 cohort was further used to identify acute phase features associated with MDC and MDC’s key chronic dimensional features by regression models. SYSU-TBI2 cohort consisted of 84 TBI patients recruited from 2021 to 2023 and was used to validate the accuracy of MDC prediction model, alongside two external cohorts, ov cohort (N=11) and Kangning cohort (N=48). SYSU-TBI2 cohort was further used, in combined with 48 additionally collected matched healthy controls, to identify in-vivo plasma proteins associated with MDC and its key chronic dimensional features.

**Findings:** Chronically, MDC-not-mild TBI patients has significantly more cognitive impairment (organic intellectual deficiency (OR=49.452 (20.366 - 148.508)), encephalomalacia (OR=3.495 (1.616 - 8.732)), radiologically evident skull defect and cranioplasty (OR=3.505 (1.937 - 6.471))), but less psychiatric impairment (postconcussional syndrome, OR=0.022 (0.006 - 0.056)) compared to MDC-mild patients (*P_FDR_* < 0.05). The best prediction model for MDC (R^2^ ranging from 0.71 to 0.79) consisted nine psychiatric-cognitive symptomatic predictors (e.g., emotional dysregulation), one radiological predictor and two predictors concerning living and social function at chronic phase, which were recognized as the 12 key chronic dimensional features underlying TBI mental prognosis measured by MDC. Acute phase brain injuries in the left hemisphere exerted a significantly stronger influence on chronic phase mental prognosis captured by the MDC than those in the right hemisphere (*P* = 3.11×10^-9^). In particular, acute phase injuries to the parietal (*P_L_vs_R_FDR_* = 6.71×10^-7^) and temporal (*P_L_vs_R_FDR_* = 4.72×10^-3^) lobes demonstrated the most marked left-right disparities, with cognitive outcomes being the most influenced aspect by such left-bias. Chronic phase Plasma GFAP, NfL, and IL-6 protein levels were significantly associated with MDC (*P_FDR_* < 0.05), with the association of NfL mediated by recovery time (nominally significant, *P_interaction_* = 0.019).

**Interpretation:** Our comprehensive evaluation of MDC as a TBI mental prognostic measurement system revealed heterogenous landscape of mental outcome of TBI. Such evaluation, along with the development of the predictive model, not only facilitated MDC’s clinical implementation but also identified potential targets for early intervention for mental complications in TBI.

**Funding:** STI2030-Major Projects, 2021ZD0202000.

## Introduction

Traumatic brain injury (TBI) affects 50 to 60 million people each year, leading to significant health deterioration and disability.^1^ The primarily cause of TBI varies across areas. In high-income regions, falls are the leading causes;^1–3^ whereas in low-income regions - despite receiving less research attention - traffic accidents account for more than half of TBI cases^1–3^ and contribute a substantial proportion of disability-adjusted life-years (DALYs) across all age groups.^4^ In long-run, TBI can lead to a range of physical, psychiatric, and cognitive consequences.^5–7^ Unfavorable chronic outcome of TBI patients are common, with an estimated 53% poor outcome rate in severe TBI patients from low-income counties.^8,9^

Stratification of TBI patients based on prognosis and identification of prognostic risk factors is important for health management but remains challenging.^1^ Prognoses of TBI are traditionally measured by Glasgow Outcome Scale (GOS) and its extended form (GOS-E), Katz Activities of Daily Living (ADL) Index, and others.^10,11^ Among those, GOS is the most well-studied measure, reflecting the overall functional recovery of TBI patients.^12^ To note, acute phase conditions do not necessarily inform chronic phase outcome. TBI patients have traditionally been classified as mild, moderate, and severe according to Glasgow Coma Scale (GCS), which is scored at acute phase.^13^ Patients classified as GCS-severe during acute phase may fully recover chronically, while those deemed GCS-mild during acute phase may experience unfavorable chronic phase outcomes.^14^ Previous studies have constructed models,^15^ such as IMPACT^16^ and IMPACT-ARACHE II,^17^ to predict TBI prognoses using information collected at acute phase, identifying prognostic risk factors such as pupillary light reactivity, epidural hematoma, and subarachnoid hemorrhage. Although these models provide good prediction accuracy, they target on the dichotomized measure of prognosis based on GOS or GOS-E, which are merely simplified assessments of the overall outcome in TBI.

One aspect that current prognostic measure and related follow-up researches are less focused on, is the mental outcome of TBI patients, despite the facts that mental disability is among the main consequences of TBI and the leading cause of DALYs among all diseases.^4^ Individuals predisposed to TBI are reported to develop somatic symptoms, cognitive impairment, and emotional or behavioral dysregulations, with a significantly increased comorbid risk with neurodegenerative diseases such as dementia^18,19^ and psychiatric disorders^20^ such as PTSD^21^ and anxiety.^22^ Radiological evidence indicates shared structural and functional brain image changes between TBI patients and patients diagnosed for psychiatric disorders or neurological diseases.^23^ In addition, neuroinflammatory biomarkers such as neurofilament-light (NfL), glial fibrillary acidic protein (GFAP), interleukin-6 (IL-6), tau, and β-amyloid (Aβ) have been reported to vary across TBI groups categorized by acute phase GCS scores.^24–27^ With all these observations, however, very few prognostic measure are specifically designed to measure the chronic mental outcome in TBI, and perhaps more unsatisfactorily, even fewer measure are designed to account for multifaceted features, ranging from symptoms to radiological changes reflecting mental disability. Symptomatic assessments provide insights into functional and behavioral impairments, yet they are usually subjective and may not reveal underlying structural or molecular changes in the brain. Conversely, radiological findings can detect objective anatomical alterations but do not necessarily correlate with cognitive or emotional functioning. Therefore, focusing on only findings from one aspect may result in an incomplete evaluation of chronic mental outcomes. Popular outcome measure of TBI such as GOS and ADL are not designed to specifically capture mental features. Quality of Life after Brain Injury (QoLIBRI) scale is designed for TBI, addressing domains such as cognition, self-perception, and emotions. However, the data gathered is entirely based on self-reports, without objective evidences such as radiological changes. To date, it remains controversial that what prognostic measure provides specific and sufficient profiling for mental outcome, and such limitation has hindered a better understanding of mental consequences in TBI.

The mental disability classification (MDC) system, commonly practiced by forensic psychiatrists in China since 1992, is specifically designed to assess chronic mental health condition in TBI patients and has the potential to address limitations mentioned above.^28^ This system was initially proposed to assess mental prognoses of TBI patients at least six months post-injury, stratifying patients into 10 grades for reasonable compensation in China. China has more TBI patients than most other countries,^9^ and the injuries are mainly caused by traffic accidents,^9^ as are those examined by the MDC system. Compared to other prognostic measures, MDC particularly focuses on psychiatric and neurological symptoms and functions, emphasizing the appearance of radiological evidence in TBI patients, while placing less emphasis on physical disability. Thus, the MDC system and its data sources would have great potential to be applied to clinical studies on mental health outcome in TBI. This necessitates a comprehensive evaluation of the critical features that the MDC system captures and the development of an accurate and practical scoring tool to enable MDC rating by the non-forensic professional, both of which have not yet been available.

In this study, we aim to systematically evaluate the MDC system as a novel prognostic measure of mental outcome in TBI by profiling the neurological, psychiatric, radiological, and molecular features it captures. We substantially improved its utility by providing an accurate and easy-to-use prediction model. Through such evaluation and application, the heterogeneous and complex nature of the chronic mental outcomes of TBI were revealed.

## Method

### Study design and participants

The SYSU-TBI is a longitudinal and observational study of patients with TBI for at least 6 months. The samples were collected at Forensic Medicine Center, Sun Yat-sen University. Two cohorts were established from this study. SYSU-TBI1 cohort consisted of 356 TBI patients recruited from 2017 to 2019. We used SYSU-TBI1 cohort to construct prediction models for MDC and perform systematic profiling of mental condition in chronic phase TBI patients. SYSU-TBI2 cohort consisted of 84 TBI patients recruited from 2021 to 2023. We used SYSU-TBI2 cohort to validate the accuracy of the prediction models of MDC, as well as investigate the association between MDC and neuroinflammatory plasma proteins. We also recruited 48 healthy controls (HC) matched by age, sex assigned at birth, and educational level to allow comparison of those proteins between TBI cases and controls. The criteria for inclusion of SYSU-TBI patients in downstream analysis included the following: 1) age from 18 to 65; 2) didn’t have a history of TBI, cerebrovascular disease, psychiatric disorder, encephalatrophy, epilepsy, or drug abuse prior to the index event; 3) have one of the following causes for the injury - mechanical external force, including traffic accident, fall, and violence, and 4) have been assessed for mental condition using MDC. For SYSU-TBI2, patients with pre-existing diseases such as hypertension and diabetes, those for whom an MDC grade could not be determined, and those self-identifying as non-Han-Chinese were excluded.

Two external cohorts, Kangning and GPMHC, were used as additional cohorts to further validate the performance of the MDC prediction models. Kangning cohort recruited 48 patients at Shenzhen Kangning Hospital (Shenzhen Mental Health Center) from 2018 to 2023. GPMHC cohort recruited 11 patients assessed for MDC at the Guangdong Provincial Mental Health Center of Guangdong Provincial People’s Hospital from 2019 to 2023.

Ethical approval was obtained from the Ethics Committee of Zhongshan School of Medicine, SYSU, Guangdong Provincial People’s Hospital, and Kangning Hospital. All phenotype data analyzed are de-identified and a waiver of informed consent was approved by the Ethics Committee aforementioned. All participants (or their legally representatives) with their plasma samples collected provided their written informed consent.

### Phenotyping

Phenotyping data were collected independently by two forensic professionals and checked for consistency. For SYSU-TBI1 cohort, we collected 189 features, including demographics, acute phase features (≤ 2 months post-TBI), chronic phase features (≥ 6 months post-TBI), and non-phase-specific features measured at time points that do not align strictly with the acute or chronic phase (Table S1). These included radiological characteristics concerning contusion and laceration, hemorrhage, cerebral ischemic infarction, skull fracture, and other aspects of brain imaging evidence. Categorical variables without an inherent order were converted into dummy variables analyzed separately in subsequent analyses. For SYSU-TBI2, GPMHC, and Kangning cohorts, we collected specific variables to validate the prediction models for MDC.

The main outcomes of this study were MDC grades assessed at least 6 months post-TBI. MDC stratifies TBI patients into 10 grades, based on the severity of long-term psychiatric and neurological symptoms, radiological evidences, BADL impairment, and social functioning impairment (Figure S1). MDC grades were determined by two forensic experts, considering various aspects such as patients’ medical records, results of mental and psychological assessment, and the examination of brain imaging and EEG (Figure S2). Apart from the MDC grades, we additionally derived an MDC severity variable with the following standard: MDC grades 1 to 6 as MDC-not-mild (grades 1-3 for MDC-severe and grades 4-6 for MDC-moderate) and MDC grades 7 to 10 as MDC-mild. The impairment of basic activities of daily living (BADL) and social function were measured by designed questionnaires (supplementary text 1) adapted from the ADL^29^ and Social Disability Screening Schedule (SDSS),^30^ categorizing TBI patients into three BADL groups and three social function groups. The GOS was dichotomized into poor (1-3) and favorable (4-5) categories, following previous practices.^31^ Diagnostic information, based on International Statistical Classification of Diseases and Related Health Problems 10th Revision (ICD-10) and Chinese classification of mental disorders (CCMD-3),^32^ was available for five diagnoses concerning psychiatric impairment (organic psychosis, organic dissociative disorder, organic mood [affective] disorders, organic personality disorder, and postconcussional syndrome) and three diagnoses concerning cognitive impairment (persistent vegetative state, organic amnesic syndrome, and organic intellectual deficiency [dementia]).

### Measure of concentrations of plasma neuroinflammatory proteins

For participants from SYSU-TBI2 cohort and matched healthy controls, blood samples were collected with EDTAK2 anticoagulant tubes (Huding, China), centrifuged within 60 minutes of collection at a low speed of 3000 rpm/min for 15 minutes, stored at -80°C, and analyzed in two batches. Plasma protein concentrations of tau, neurofilament-light (NfL), glial fibrillary acidic protein (GFAP), ubiquitin carboxyl-terminal hydrolase L1 (UCH-L1), interleukin 6 (IL-6), Aβ40, and Aβ42 were measured using Single-molecule Array (Simoa) N4PA, N3PA, and IL-6 Advantage Kits (Quanterix, Billerica, USA).

### Construction and validation of prediction models for MDC

Two machine learning algorithms, Least Absolute Shrinkage and Selection Operator (LASSO) and random forest (RF), were used to screen variables and build prediction models for MDC using the training set (the SYSU-TBI1 cohort, N=356). The effectiveness of the final model was verified in three test cohorts, SYSU-TBI2 (N=84), Kangning (N=48) and GPMHC (N=11). In the training cohort, to avoid severe sample loss while remaining as much features as possible, we only remained features with a missing sample proportion of less than 25% (N_total features_=189, N_passed features_=181, eight features were filtered out in this step). The 181 features included those measured at different phases and demographic variables (Table S1). Missing values in the remained features were then imputed with the RF algorithm implemented through the “rfImpute” function of the “randomForest” package. The iteration parameter *iter* for imputing missing value was set to 6.

Feature selection: we performed feature selection using the LASSO and RF methods. In brief, to ensure model stability we generated 200 seeds, with each seed randomly selecting a subset of 80% of the samples from the training set. For each subset, we applied 1000 times 10- fold cross validation using LASSO approach to identify features that appeared in more than 10% of the models. These features were then supplied to LASSO to build a subset-specific model. In total of 200 subset-specific models were constructed, from which we identified 12 features that were included in more than 75% of the subset-specific models. We then used the 12 features and full samples in the training set to randomly construct LASSO prediction models 1,000 times and found that the 12 features have an occurrence rate of 100%. We used the 12 features to build random forest models and then select features with variable contributions greater than 0, in this step, we tested all possible parameter combinations of *mtry* (1, 2, 3, 4, 5, 6), *nodesize* (2, 3, 4, 5), *nsplit* (1, 2, 3, 4, 5), and *ntree* (200, 300, 400, 500, 600, 700, 800, 900, 1000) and employed five-fold cross validation to identify the parameter combination with the largest R^2^. We found all the 12 features in the random forest model had variable contributions greater than 0. All of the 12 features were chronic-phase features and were selected and included in final model construction procedures below.

To build the final LASSO model for MDC, we used the 12 selected features, full training samples and 10-fold cross validation with the parameter *type.measure = “mse”* (minimizing mean squared error).

To build the final RF model, we tested all possible parameter combinations of *mtry* (1, 2, 3, 4, 5, 6), *nodesize* (2, 3, 4, 5), *nsplit* (1, 2, 3, 4, 5), and *ntree* (200, 300, 400, 500, 600, 700, 800, 900, 1000). We employed five-fold cross validation to identify the parameter combination with the largest R^2^. Then we used the parameter combination with the largest R^2^ and all training samples to build the final random forest model.

### Construction and validation of prediction models for BADL

Two machine learning algorithms, LASSO and RF, were used to screen variables and build prediction models for BADL impairment using the training set (the SYSU-TBI1 cohort, N=353, three cases were removed due to missing BADL information). The effectiveness of the final model was verified in the SYSU-TBI2 cohort with BADL model selected features (N=36). In the training set, input features included 177 variables including those measured at different phases and demographic variables. MDC, degree of BADL impairment, social functioning impairment, and degree of social functioning impairment were not included.

Feature selection: we performed feature selection using the same steps as the analysis for MDC. We identified the parameter combination with the largest area under curve (AUC). 13 features were selected and included in final model construction procedures below.

To build the final LASSO model for BADL impairment, we used the selected features and full training samples, and we chose 10-fold cross validation and the parameter *type.measure=“class”* (minimizing misclassification error).

To build the final RF model, we tested all possible parameter combinations of *mtry* (1, 2, 3, 4, 5, 6), *nodesize* (2, 3, 4, 5), *nsplit* (1, 2, 3, 4, 5), and *ntree* (200, 300, 400, 500, 600, 700, 800, 900, 1000). We employed five-fold cross validation to identify the parameter combination with the largest AUC. Then we used the parameter combination with the largest AUC and all training samples to build the final random forest model.

### Statistical analyses

Cohort characteristics were compared using Mann-Whitney U test for continuous characteristics and χ² tests for comparison of categorical characteristics. Fisher’s exact tests were used to test for the enrichment of diagnoses, BADL, and social functioning impairment at chronic phase, as well as radiological features at both acute and chronic phases, in MDC severity groups using SYSU-TBI1 cohort. Associations between MDC-group-varied radiological features and diagnoses/symptoms were tested using logistic regression. Months post-injury was the only covariate jointly fitted when analyzing the association between acute and chronic phase radiological features, as none of the demographic characteristics (age, sex assigned at birth, ethnicity, educational level, family history of mental illness, history of diabetes, history of gout, and history of hypertension) was significantly associated with the chronic phase radiological characteristics in corresponding simple regression analysis (*P_FDR_*>0.05). Multiple testing was corrected using the false discovery rate (FDR) method.

The association tests between acute phase features (as dependent variable, N_features_=86) and MDC or the key chronic dimensional features of MDC (as independent variable, N_features_=11, “unconsciousness” was excluded because of a low positive rate (0.6%) in SYSU-TBI1 cohort) were performed using logistic (for binary and dummy independent variable) or linear (for continuous and ordinal independent variable) regression. Continuous and ordinal variables were scaled to a range of 0 to 1. No additional covariates were jointly fitted because we examined eight demographic features (age, sex assigned at birth, ethnicity, educational level, family history of mental illness, history of diabetes, history of gout, and history of hypertension) and months post-injury and found they were not significantly associated (*P_FDR_*>0.05) with MDC and its key chronic dimensional features. Multiple testing correction was performed using the FDR method.

The associations of plasma protein concentration with the TBI exposure (TBI vs control), with the MDC, and with MDC’s key chronic dimensional features was examined using linear regression in the SYSU-TBI2 cohort (N_TBI_=84, with matched controls, N_control_=48). Plasma protein concentrations were Z-score normalized (mean = 0, standard deviation = 1) using R. The demographic characteristics were comparable between mild and not mild MDC groups in the SYSU-TBI2 cohort (Table S2). Age, sex assigned at birth, educational level, months post-injury, and analysis batches were jointly fitted as covariates. *P* values were adjusted using the FDR method. All analyses were done with R (version 4.3.1).

## Results

After applying stringent inclusion and exclusion criteria, 499 chronic TBI (≥6 months post TBI) cases were available across four cohorts (SYSU-TBI1, SYSU-TBI2, Kangning, GPMHC) from three centers. Of these, 84 cases from the SYSU-TBI2 cohort had plasma concentrations measured for seven proteins (Figure 1A, Table S2). The SYSU-TBI1 cohort (N=356) was treated as the training dataset when constructing the prediction model for MDC; whilst the SYSU-TBI2 cohort (N=84) was used as an independent test dataset from the same center. The Kangning and the GPMHC cohorts were used as two external test datasets to further validate the MDC model. In the training dataset, the ratio of male and female was 75.8%:24.2%, the mean age was 43.1(SD±12.8), and 80.3% TBI patients had an educational attainment of junior high school or below (Table S3). Comparing to other chronic outcome measures of TBI, MDC is specifically designed for TBI patients and primarily focuses on symptoms concerning both psychiatric and cognitive impairment and radiological evidence present in patients more than 6 months post-TBI (Figure 1B).

**Figure 1.**
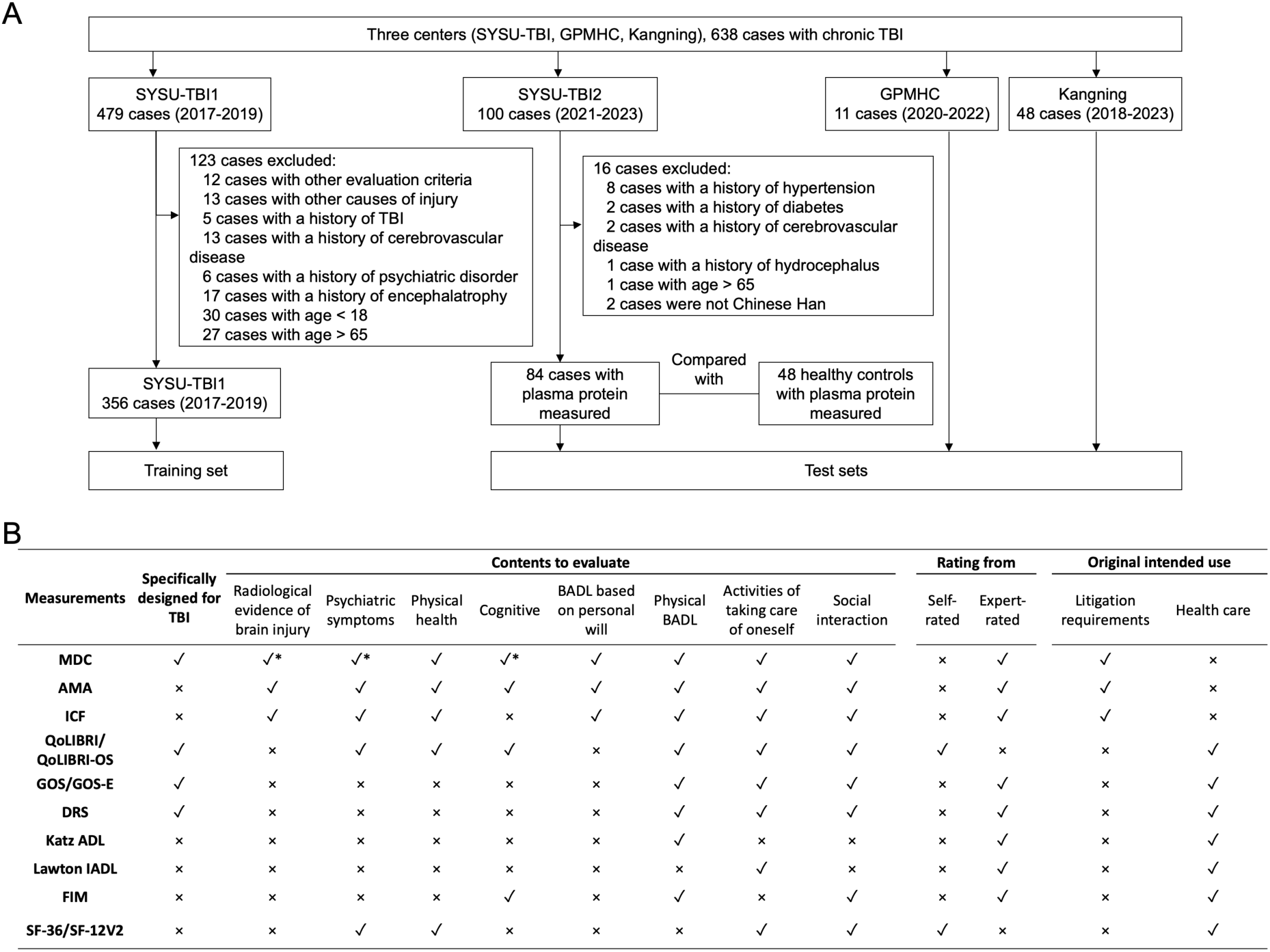
Recruitment flowchart and the comparison between mental disability classification and other measures of chronic phase TBI outcome. A: Recruitment flowchart. B. Mental disability classification and other measures of chronic phase TBI outcome. ✓: The aspect is considered in this measure; ×: The aspect is not considered in this measure; *: The aspect is given substantial weight in MDC.

In the MDC system, a lower grade corresponds to a poorer mental condition (Figure S1). According to the MDC rating, mental outcome of TBI is highly heterogeneous, patients rated with the same GCS level during the acute phase substantially differed in their MDC grades rated at the chronic phase (Figure 2A). When grouping TBI patients into chronically MDC-mild (MDC grades 7-10) and MDC-not-mild (MDC grades 1-6) categories, a significantly lower occurrence of psychiatric disorder such as postconcussional symptoms at chronic phase was observed in the MDC-non-mild group (*P_FDR_*=8.77×10^-23^, occurrence rate: 79% (mild) vs 8% (not-mild), OR= 0.022 (0.006 - 0.056). Figure 2B, S3, Table S4). For the same group (MDC-not-mild), in contrast, a significant rise of cognitive impairments, such as intellectual deficiency (dementia), was detected at chronic phase (*P_FDR_* =6.94×10^-26^, occurrence rate 16% (mild) vs 90% (not-mild), OR= 49.452 (20.366 - 148.508), Figure 2B, Table S4). Most TBI patients demonstrated chronically social functioning impairment, but the MDC-not-mild group developed significantly more BADL impairment than the mild group (*P_FDR_* =5.59×10^-22^, occurrence rate :34% (mild) vs 100% (not-mild), Figure 2B, Table S4). Based on the radiological evidence present at chronic phase, the MDC-not-mild group underwent significantly more skull defect or cranioplasties (*P_FDR_* =4.21×10^-5^, occurrence rate: 31% (mild) vs 61% (not-mild), OR= 3.505 (1.937 - 6.471)), and displayed a significantly higher incidence of encephalomalacia compared to the MDC-mild group (occurrence rate: 64% (mild) vs 85% (not-mild), OR= 3.495 (1.616 - 8.732), *P_FDR_*=1.94×10^-3^, Figure 2C, Table S4). The appearance of skull defect or cranioplasties and encephalomalacia in chronic phase are positively associated with several radiologically-evident impairments in acute phase (*P_FDR_* < 0.05), examples included the positive association between acute phase – brain herniation and chronic phase – skull defect or cranioplasty (Table S4, Figure 2D). Additionally, chronic phase skull defect or cranioplasties and encephalomalacia are also positively associated with chronic phase intellectual impairment and negatively associated with chronic phase psychiatric disorders (*P_FDR_* < 0.05, Figure 2D).

**Figure 2.**
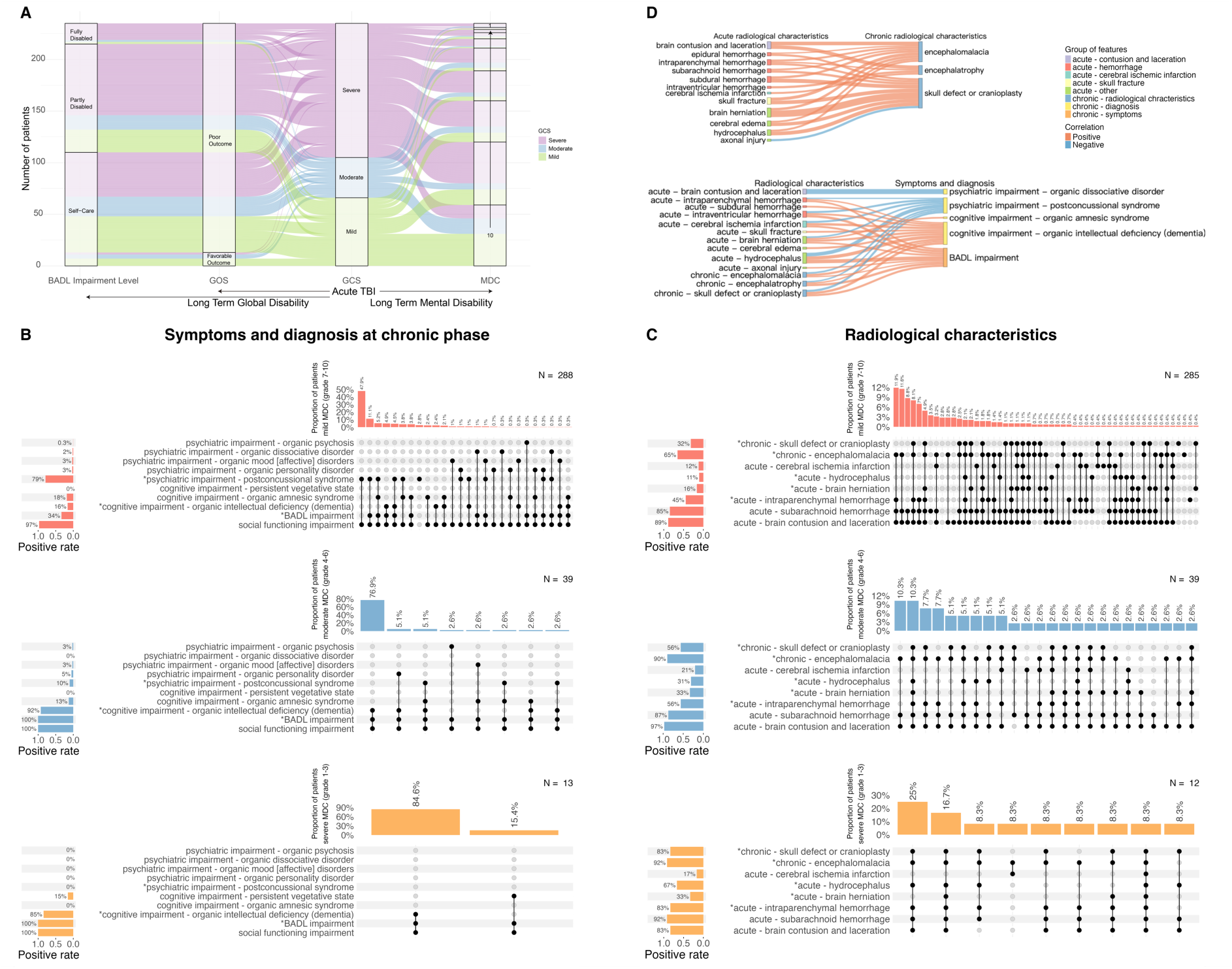
Features of patients from different MDC groups. **A.** TBI patients from SYSU-TBI1 cohort belonging to different acute phase GCS groups categorized by GCS scores, and chronic phase groups categorized by MDC, GOS and BADL. GCS: Glasgow Coma Scale, MDC: Mental Disability Classification, GOS: Glasgow Outcome Scale, BADL: Basic Activities of Daily Life. Arrows represent the direction of time flow. **B.** Chronic-phase symptoms and diagnosis in TBI patients with different MDC severities. Mild MDC: MDC grades 7-10, moderate MDC: MDC grades 4-6, severe MDC: MDC grades 1-3. **C.** Acute- and chronic-phase radiological characteristics in TBI patients with different MDC severities. * in B&C: significant difference (*P_FDR_*<0.05) across MDC severity groups by Fisher’s exact test. **D.** Significant associations between radiological characteristics at either the acute or chronic phase and chronic phase symptoms and diagnosis.

We built prediction models for MDC using LASSO and RF using SYSU-TBI1 cohort as the training dataset and the rest three cohorts as the test datasets, which not only effectively predicted MDC (best performance is by RF method: R^2^_test-SYSU-TBI2_=0.788, R^2^_test-Kangning_=0.744, R^2^_test-GPMHC_= 0.711), but also deconstructed the MDC into critical symptomatic and radiological dimensions present at the chronic phase (Figure 3A). The models constructed by both LASSO and RF methods include the same set of 12 chronic phase features, 9 of which are psychiatric and neurological symptoms (e.g., *degree of insight impairment*), 2 concern impairments of BADL and social function, and one is a radiological feature (*number of lobes with encephalomalacia*). These features could be considered as the key chronic dimensional features underlying the MDC. According to the inclusion mean squared error (MSE) of the model with the best performance (the RF model), the top three most important dimensional features are *degree of insight impairment, hypobulia or hyperbulia, and emotional dysregulation*. We additionally build prediction models for BADL impairment as the outcome. Different from MDC, the key dimensions of BADL impairment consisted of both acute and chronic phase features including three physical characteristics (Figure 3B). Overall, whilst traditional TBI outcome measures such as ADL assigned substantial weight on physical characteristics, MDC mainly captured multi-level information involving psychiatric, neurological symptoms and radiological changes.

**Figure 3.**
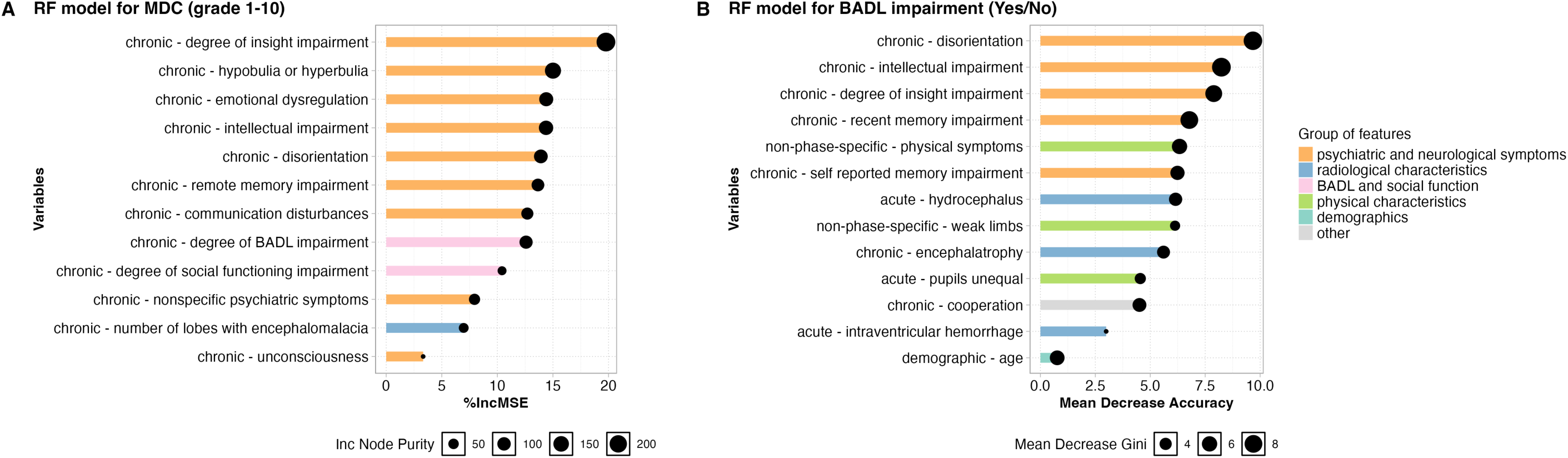
The variable importance of the prediction models for TBI chronic outcomes. **A.** The variables importance of the RF prediction model for MDC. **B.** The variable importance of the RF prediction model for BADL impairment. RF: Random Forests; MDC: Mental Disability Classification; BADL: Basic Activities of Daily Life.

We wondered whether the chronic phase mental outcome assessed by the MDC can be linked to acute phase features. We first considered the potential impact from basic features, including eight demographic variables (age, sex assigned at birth, ethnicity, educational level, family history of mental illness, history of diabetes, history of gout, and history of hypertension) and months post-injury, by performing association tests between each of the those features and MDC, and detected no significant association (*P_FDR_* > 0.05). We then analyzed the association between MDC and 86 acute features, revealing that 34 acute features are significantly associated with MDC (*P_FDR_* < 0.05, Figure 4A). These acute features can be categorized to seven groups (e.g. psychiatric and neurological symptoms, contusion and laceration, and hemorrhage, Figure 4A). The most associated acute features with MDC are coma duration and hydrocephalus (*β*_coma_duration_= -3.088, *P_FDR_*__coma_duration_= 3.17×10^-14^; *β*_hydrocephalus_ = -2.392, *P_FDR_*__hydrocephalus_= 3.90×10^-15^, Table S5).

**Figure 4.**
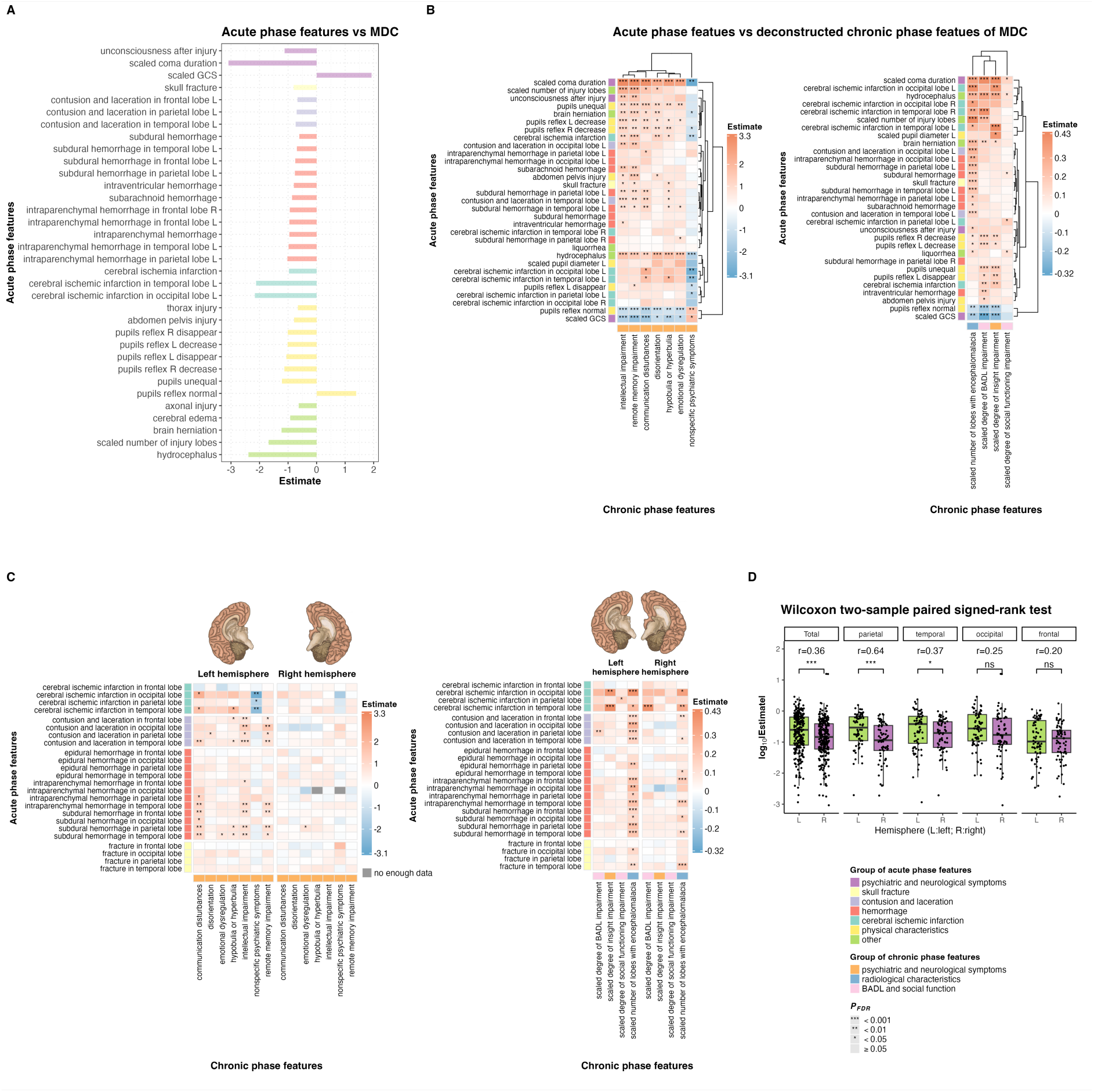
The associations of the chronic phase MDC and its key dimensional features with acute phase features in TBI patients. L: left hemisphere. R: right hemisphere. Estimate: regression coefficient. **A.** Acute phase features significantly associated with the MDC. MDC: Mental Disability Classification. **B.** The most significant associations between acute phase features and MDC’s deconstructed chronic dimensional features (the top 10 most significantly associated acute phase features for each chronic phase feature were displayed). **C.** Association results between the key chronic dimensional features of MDC and radiologically evident brain injuries in left and right hemispheres at acute phase. **D.** Results of Wilcoxon tests comparing the absolute value of regression coefficient between acute phase injury in left and right hemispheres. r: effect size, log_10_|Estimate|: the base-10 logarithm of the absolute regression coefficient. ***: *P_FDR_*<0.0001, **: *P_FDR_*<0.001, *: *P_FDR_*<0.05, ns: *P_FDR_*≥0.05.

We next sought to investigate specific connections between acute phase features and the key chronic phase psychiatric, neurological, and radiological aspects underlying MDC (Figure 4, Table S6). This was realized by analyzing the association between 86 acute phase features and the 11 deconstructed chronic dimensional features of the MDC model (one deconstructed chronic phase feature, unconsciousness, has less than 1% positive rate so it was not used in this analysis. Figure 4B, Figure S4). The results showed that coma duration and hydrocephalus, the acute phase features with the strongest association with MDC, also displayed the most widespread associations with deconstructed MDC features. Remote memory impairment was the most associated neuropsychiatric symptom at chronic phase with acute phase features (associated with coma duration, OR = 26.550 (10.271 - 73.617), *P_FDR_* = 1.40×10^-8^, Table S6). Intriguingly, a systematic left-hemisphere bias was observed when examining the association between acute phase radiological evidences of brain injury and MDC’s key chronic dimensional features (*P_Wilcoxon_*=5.36×10^-10^. Figure 4C & D). Multiple radiologically evident acute phase factors, concerning contusion and laceration, subdural hemorrhage, cerebral ischemic infarction, and intraparenchymal hemorrhage, displayed stronger associations with MDC’s key chronic dimensional features when the injury occurred in the left hemisphere compared to the right (Figure 4C). To note, acute phase injuries in parietal and temporal lobes were particularly prominent for this left-bias *(P_FDR_* < 0.05, r > 0.35. Figure 4D), and cognitive MDC features such as remote memory impairment were more influenced by this bias (Figure S5). The top three important dimensional features of MDC (Figure 3A) were significantly associated with a number of acute phase injuries in the left hemisphere of the temporal lobe, but not the right hemisphere counterparts. For example, acute phase cerebral ischemic infarction in the left-hemisphere temporal lobe, but not on the right side, increased the risk for higher *degree of insight impairment* at chronic phase (*β* = 0.422, *P_FDR_* = 5.04×10^-4^); acute phase subdural hemorrhage, cerebral ischemic infarction and contusion and laceration in the left-hemisphere temporal lobe, but not on the right side, are high-risk factors for chronic phase – *hypobulia or hyperbulia* (*P_FDR_* < 0.05); acute phase subdural hemorrhage in the left-hemisphere temporal lobe, but not on the right side, increases the risk for chronic - *emotional dysregulation* (OR = 1.983 (1.211 - 3.244), *P_FDR =_* 3.25×10^-2^). Such left-hemisphere-bias was likely due to the dominant hemisphere effect, as we observed an even more prominent bias in all lobes when comparing dominant vs non-dominant hemisphere (Figure S6).

Finally, we investigated the molecular signatures of the chronic phase mental outcome of TBI, as reflected by the MDC system, by performing association tests between plasma levels of seven neuroinflammatory proteins and MDC, and MDC’s chronic dimensional features. We used multiple linear regression with age, sex assigned at birth, educational level, months post-injury, and batches jointly fitted as covariates. The results showed that plasma concentration of GFAP, NfL, IL-6, Aβ40 and Aβ42 were significantly elevated at the chronic phase in TBI patients compared to the HC group (*P*_FDR_ < 0.05, Figure 5A), while Tau and UCH-L1 were not (*P*_FDR_ > 0.05, Figure S7A-B). Within the TBI group, plasma levels of GFAP, NfL and IL-6 showed significant associations with MDC in regression models where outcome variable was either the 10-level MDC grades or the two-level MDC severity groups (*P*_FDR_< 0.05, Figure 5B-C), while no significant association was detected for other plasma proteins (*P*_FDR_ > 0.05, Figure S7C-K). To account for the potential influence of somatic injuries on observed associations for MDC, we performed sensitivity analysis by adding somatic injury as a covariate. The association of Plasma NfL, GFAP and IL-6 with MDC remained significant, although a decrease of effect size from NfL was observed, indicating that at least part of plasma protein changes was independently related to mental impairment (Table S7).

**Figure 5.**
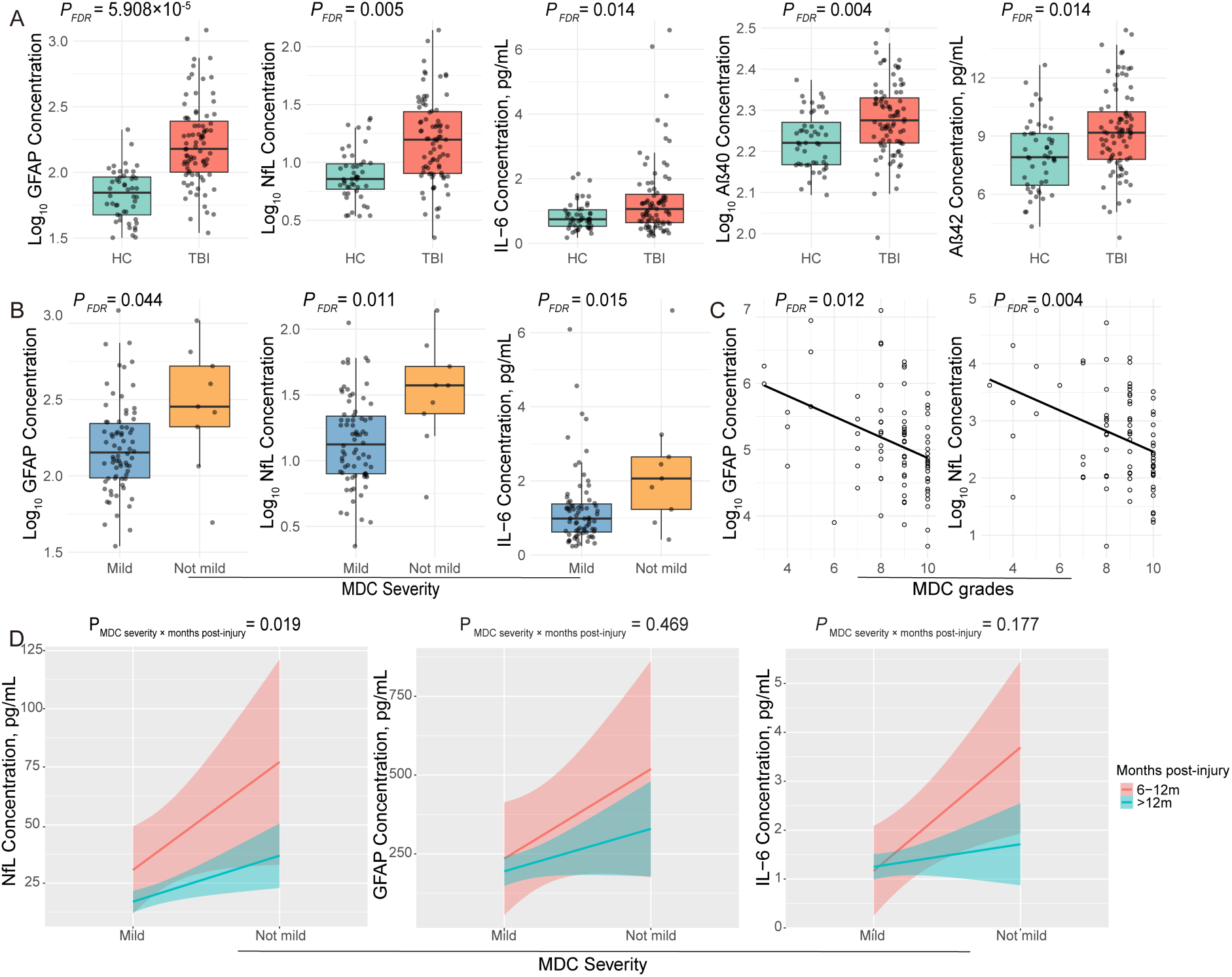
Plasma proteins significantly associated with mental prognosis of TBI. A: Significantly differentiated protein concentration between TBI and HC groups; B and C: Significantly differentiated protein concentration amongst TBI patients of varied MDC; D: Interactive effect between MDC severity and months post-injury on NfL, GFAP and IL-6. P-values shown in figure 5D are all raw *P* values. Months post-injury was treated as a continuous variable in interactive effect analyses.

We further explored specific links between MDC-associated proteins with the key chronic dimensional features underlying MDC (Table S8). NfL and GFAP concentrations were both significantly correlated with number of lobes with encephalomalacia, disorientation, intellectual impairments, and degree of BADL impairment levels. Additionally, NfL was uniquely significantly correlated with communication disturbances and nonspecific psychiatric symptoms, and GFAP was uniquely significantly correlated with hypobulia or hyperbulia and degrees of insight impairment (Table S8). As IL-6 was significantly associated with the two-level MDC severity groups, consistently, it was also significantly associated with degrees of insight impairment and disorientation (Table S9), two key chronic dimensional features of the optimal model for the two-level MDC severity groups (Figure S8).

We also compared the protein association patterns for the MDC, the outcome measure of mental health; and the BADL, the outcome measure of overall functional disability. While GFAP and NfL showed significant associations with both MDC and BADL, IL6 was uniquely associated with MDC (Figure 5B-C). In addition, a nominally significant interactive effect between MDC severity and recovery time was detected on NfL (*P*_interaction_ =0.019, *P*_FDR-interaction_=0.056) but not on other proteins (Figure 5D), indicating that patients recruited at an earlier time point following TBI displayed a stronger correlation between MDC severity and NfL than those recruited at later time. Other proteins also displayed similar trend of recovery-time-stratified associations with MDC (Figure 5D). A similar interaction effect was observed when examining BADL and months post-injury in relation to NfL (Figure S9, *P*_interaction_=0.011).

## Discussion

We conducted the first systematic evaluation of the MDC system and unveiled the heterogenous landscape of chronic mental health consequences of TBI. At chronic phase, patients rated as mild by MDC exhibited a higher prevalence of psychiatric impairment, while those rated as more severe by MDC developed significantly more cognitive impairment, along with a higher prevalence of radiologically evident skull defect or cranioplasty and encephalomalacia. Using machine learning methods, our MDC prediction models substantially enhanced MDC’s utility and identified nine psychiatric-cognitive symptoms, one radiological feature, and two measures of impairments in activities of daily living and social function at the chronic phase as the key dimensional features underlying MDC. The association tests between acute vs chronic phase features underscored the much stronger influence from brain injuries in the left hemisphere on mental outcome of TBI as compared to the right hemisphere, with injuries in the parietal and temporal lobes showing the most pronounced left-right differentiated effect. Plasma GFAP, NfL, and IL-6 protein levels are significantly associated with MDC, highlighting their potential as biomarkers for chronic mental outcome in TBI.

The heterogeneity of TBI mental outcome is reflected in the spectrum of MDC grades observed at the chronic phase even for TBI patients rated as similar injury level based on GCS scores at the acute phase (Figure 2A). In our training cohort (SYSU-TBI1), 39.9% of patients were classified as GCS-severe in the acute phase, which is significantly higher than the 21% observed in hospital-sourced cohorts^26^. This reflects an enrichment of severe cases in forensic-source cohorts where chronic phase organic impairment is prerequisite for forensic examination. Consistently, these judicial cases, whether rated as acute phase-GCS-mild, - moderate or -severe, were mostly categorized as having a chronic poor outcome based on the traditional TBI outcome measure GOS (276/293=94.2%. Table S3). The chronic mental condition of judicial patients, however, were mostly rated to be MDC-mild (7-10 grades, 83.7%, Table S3). In general, the severity of injury evaluated at the acute phase was positively correlated with the degree of mental impairment observed at the chronic phase, as a higher proportion of acute phase-GCS-mild cases were rated as chronic phase-MDC-mild (92.3%) compared with acute phase-GCS-severe cases (78.2%). However, this also indicated that 7.7% acute phase-GCS-mild cases progressed to severe mental impairment at the chronic phase, and on the other hand, 11.8% acute phase-GCS-severe cases chronically recovered to a mental state comparable to that of most acute phase GCS-mild cases. Our results also indicated one form of psychiatric impairment, the postconcussional syndrome, can persist long for large proportion of MDC-mild cases (Figure 2B), aligning with previous observations from hospital-sourced TBI cases ^33^. On the contrary, MDC-severe cases rarely developed postconcussional syndrome, but instead, exhibited high rate of organic intellectual deficiency. This echoes previously reports on the inverse relationship between psychiatric symptom severity and TBI severity,^34^ likely attributable to the impaired insight and the resulted reduced emotional distress or apathy associated with severe TBI.^35^

We further approached the heterogeneous TBI mental outcome through deconvoluting the MDC grades into 12 key chronic dimensional features. The identified key chronic phase dimensions include both psychiatric-cognitive symptoms (e.g. *degree of insight impairment, hypobulia or hyperbulia, and emotional dysregulation*) and radiological feature (*number of lobes with encephalomalacia*), highlighting the MDC system’s strength in integrating multi-level evidence of both subjective and objective, symptomatic and radiological. The deconvolution strategy, combined with the longitudinal design of the SYSU-TBI1 cohort that encompasses both acute- and chronic-phase variables, enabled the identification of shared and non-shared acute phase risk factors not only for the MDC itself but also for specific key chronic dimensions underlying the MDC. The most shared acute phase risk factors include coma duration and hydrocephalus, the two known risk factors of poorer TBI prognosis,^36,37^ as they display widespread associations with deconstructed MDC dimensions. What we additionally revealed here, was that it is the chronic phase – remote memory impairment to be the most influenced chronic neuropsychiatric dimension by a longer acute phase coma duration. We also identified specific acute-chronic links such as the positive association between acute phase amnesia after injury and chronic-phase remote memory impairment (Table S6), as no other chronic phase features significantly associated with this acute phase feature. These findings uncovered the early origins of the heterogenous mental condition at chronic TBI phase and provided specific targets for early risk assessment and intervention to improve mental prognosis of TBI patients.

We found that the acute phase features of brain injury in left hemisphere, which is the dominant hemisphere for most participants (98% TBI patients in SYSU-TBI1 are self-identifying right-handed), are more strongly associated with the key chronic dimensional features of MDC than the counterpart in right hemisphere. Injuries in parietal and temporal lobe of the left hemisphere are particularly influential for the chronic features. Previous studies have reported that parietal and temporal lobes regulate memory, communication functions.^38–41^ Our result indicated that acute phase brain injury in the left hemisphere of those lobes, rather than the right side, are significantly associated with remote memory impairment, communication disturbances, and intellectual impairment in the chronic phase of TBI, all of which are the key chronic dimensional features of the MDC. These results align with previous researches that have identified associations between parietal^40,41^ or temporal lobes^42–44^ and those functions, highlighting the need of special attention on these aspects of long-term mental outcome in TBI patients with acute phase injury to the parietal and temporal cortices of their dominant hemisphere.

Beyond aiding the deconvolution of MDC, our prediction model for MDC enhanced objectivity and utility of the MDC system in assessing chronic mental outcomes of TBI patients. The model simplified the complex rating processes that are previously rely on forensic psychiatrists’ expertise (Figure S1,2) into 12 easily measurable variables at the chronic phase, and display a good predictive accuracy validated across cohorts. This ensured the MDC system’s generalizability for broader application, introducing MDC’s promising potential utility beyond forensic settings in China to clinical researchers aiming to disentangle the chronic mental health of TBI patients worldwide. To note, our findings highlighted several advantages of forensic-sourced TBI biobank in researches of chronic TBI. These included the rich phenotyping information on chronic mental health of patients which can be obtained through the forensic examination, in contrast to the relatively limited phenotyping information available from hospital- or general population-sourced follow-up samples. Additionally, forensic biobanks of our type offer an inherent longitudinal data source with both acute phase and chronic-phase information available, as acute phase medical records are collected during chronic-phase forensic assessments. Bridging the gap between metrices from forensic and hospital-practices needs courage but is full of opportunities. Our study offers a comprehensive evaluation paradigm to help close this gap.

Our results from association tests of plasma proteins supported the biological relevance of the TBI mental outcome measured by the MDC system. We first compared chronic TBI cases to controls and detected significantly increased plasma GFAP, NfL and IL-6 concentrations in chronic TBI group (Figure 5A), which are consistent with previous studies^24,26,45^. Concentrations of Aβ40 and Aβ42 were also increased in chronic TBI patients group (Figure 5A), aligning with epidemiological observations that TBI predisposes risk for neurodegenerative diseases^46^. No significant change was detected in UCH-L1 concentrations, suggesting that the UCH-L1 response post TBI is likely to confine to acute phase^24,26^. Plasma tau concentrations did not show significant changes, which doesn’t align with some of the previous observations^24,47^, possibly due to ethnic differences and limitation of sample size. We then compared protein concentrations amongst TBI patients and detected significant and positive associations of plasma GFAP, NfL, and IL-6 with the severity of chronic mental disability as reflected by MDC, implicating the potential of these proteins as biomarkers for long-term neurological and psychiatric outcomes (Figure 5B). Additionally, we found that these plasma proteins were only associated with a subset of chronic dimensions of MDC (Table S8-10), implicating diverse molecular mechanism underlying different aspects of TBI mental outcome and as a result, the necessity of specific biomarkers in future researches. Our study is one of the very few studies to investigate chronic-phase plasma protein difference amongst TBI patient groups categorized by chronic mental outcomes, as opposed to most prior studies that have compared acute- or chronic-phase measured proteins in TBI groups defined by GCS levels at acute phase^24,26,45^. Thus, our study serves as an important complement to existing studies by identifying candidate chronic-phase biomarkers for mental outcomes in TBI.

To note, the correlation between chronic-phase NfL and mental outcome (MDC) decreased overtime, other plasma proteins also displayed similar trend (Figure 5D). Previously, Wong et al. suggested that NfL could originate from both brain and peripheral injuries, but the concentration changes do not affect NfL’s specificity as a biomarker for moderate TBI, while the performance may decrease for less severe TBI cases^48^. Our findings on human samples aligned with the finding on mice. NfL’s association with MDC was attenuated after conditioning on somatic injury, though its correlation with MDC persists. In contrast, GFAP, though also expressed in the periphery^49,50^, showed little change in its association with MDC after conditioning on somatic injury. These results indicated that evaluation of chronic mental outcome biomarkers such as NfL should take recovering time and somatic injury into account.

Our study has several limitations. Although the sample size of our training cohort is among the largest of forensic-sourced TBI cohorts to our knowledge, the sample size of validation analyses is relatively limited. Future studies validating the MDC model in larger samples are needed. Despite our study being one of the few that bridge the acute and chronic phases mental-related features of TBI, we only have data from two time points with a long interval time (on average 13 months). Future studies with data collected at multiple time points will enable the analyses of symptom progression, radiological changes, and biomarker trajectories during TBI recovery process. Due to the limited availability of other TBI outcome measures in our samples, we only compared MDC with BADL and dichotomized GOS. A more comprehensive collection of TBI outcome measures will be necessary to identify unique and shared information captured in different TBI measures. Finally, we only tested 7 proteins for the association with MDC, future studies examining molecular features genome-wide with MDC will provide a better molecular profiling of the mental outcome in TBI.

In summary, mental prognosis in TBI is highly heterogenous and can be quantified using the MDC system. Deconvolution of MDC by constructing prediction models revealed key chronic dimensions concerning psychiatric-cognitive symptoms and radiological features, which are associated with shared and specific acute phase risk factors, as well as chronic-phase neuroinflammatory plasma proteins. Such systematic evaluation of the MDC system and the construction of its prediction model facilitated the application of MDC and provided targets for early interventions for TBI patients susceptible for poor mental prognosis.

### Data Sharing Statements

The data presented in this study are under controlled access due to data privacy laws related to patient consent for data sharing. Application should be made to.

## Data Availability

The data presented in this study are under controlled access due to data privacy laws for data sharing. Application should be made to corresponding authors.

## Acknowledgement

We acknowledge supports from the STI2030-Major Projects, 2021ZD0202000 (YZ, XY).

## References

1. Maas AI, Menon DK, Manley GT, et al. Traumatic brain injury: progress and challenges in prevention, clinical care, and research. The Lancet Neurology 2022.

2. Ghneim M, Brasel K, Vesselinov R, et al. Traumatic Brain Injury in Older Adults: Characteristics, Outcomes, and Considerations. Results From the American Association for the Surgery of Trauma Geriatric Traumatic Brain Injury (GERI-TBI) Multicenter Trial. Journal of the American Medical Directors Association 2022; 23(4): 568–75.e1.

3. Chikhladze NN, Tsiskaridze A. Epidemiological features of traumatic brain injuries from a first level trauma care national medical center in Georgia. One Health and Risk Management 2023; 2(4): 5–11.

4. Ferrari AJ, Santomauro DF, Aali A, et al. Global incidence, prevalence, years lived with disability (YLDs), disability-adjusted life-years (DALYs), and healthy life expectancy (HALE) for 371 diseases and injuries in 204 countries and territories and 811 subnational locations, 1990–2021: a systematic analysis for the Global Burden of Disease Study 2021. The Lancet 2024; 403(10440): 2133–61.

5. Agtarap SD, Campbell-Sills L, Jain S, et al. Satisfaction with life after mild traumatic brain injury: a TRACK-TBI study. Journal of neurotrauma 2021; 38(5): 546–54.

6. Pavlovic D, Pekic S, Stojanovic M, Popovic V. Traumatic brain injury: neuropathological, neurocognitive and neurobehavioral sequelae. Pituitary 2019; 22(3): 270–82.

7. Van Reekum R, Bolago I, Finlayson M, Garner S, Links P. Psychiatric disorders after traumatic brain injury. Brain injury 1996; 10(5): 319–28.

8. Gong R, Liang Y, Gao G, Bao Y. Chinese head trauma data bank: factors of short-term prognosis. Chin J Neurosurg 2014; 30: 56–8.

9. Jiang J-Y, Gao G-Y, Feng J-F, et al. Traumatic brain injury in China. The Lancet Neurology 2019; 18(3): 286–95.

10. Jennett B, Bond M. Assessment of outcome after severe brain damage: a practical scale. The Lancet 1975; 305(7905): 480–4.

11. Wilson JL, Pettigrew LE, Teasdale GM. Structured interviews for the Glasgow Outcome Scale and the extended Glasgow Outcome Scale: guidelines for their use. Journal of neurotrauma 1998; 15(8): 573–85.

12. McMillan T, Wilson L, Ponsford J, Levin H, Teasdale G, Bond M. The Glasgow Outcome Scale - 40 years of application and refinement. Nat Rev Neurol 2016; 12(8): 477–85.

13. Teasdale G, Jennett B. ASSESSMENT OF COMA AND IMPAIRED CONSCIOUSNESS: A Practical Scale. The Lancet 1974; 304(7872): 81–4.

14. Tenovuo O, Diaz-Arrastia R, Goldstein LE, Sharp DJ, van der Naalt J, Zasler ND. Assessing the Severity of Traumatic Brain Injury—Time for a Change? Journal of Clinical Medicine 2021; 10(1): 148.

15. Matsuo K, Aihara H, Nakai T, Morishita A, Tohma Y, Kohmura E. Machine Learning to Predict In-Hospital Morbidity and Mortality after Traumatic Brain Injury. J Neurotrauma 2020; 37(1): 202–10.

16. Steyerberg EW, Mushkudiani N, Perel P, et al. Predicting outcome after traumatic brain injury: development and international validation of prognostic scores based on admission characteristics. PLoS medicine 2008; 5(8): e165.

17. Raj R, Siironen J, Kivisaari R, Hernesniemi J, Skrifvars MB. Predicting outcome after traumatic brain injury: development of prognostic scores based on the IMPACT and the APACHE II. J Neurotrauma 2014; 31(20): 1721–32.

18. Barnes DE, Kaup A, Kirby KA, Byers AL, Diaz-Arrastia R, Yaffe K. Traumatic brain injury and risk of dementia in older veterans. Neurology 2014; 83(4): 312–9.

19. Gardner RC, Burke JF, Nettiksimmons J, Kaup A, Barnes DE, Yaffe K. Dementia Risk After Traumatic Brain Injury vs Nonbrain Trauma: The Role of Age and Severity. JAMA Neurology 2014; 71(12): 1490–7.

20. Koponen S, Taiminen T, Portin R, et al. Axis I and II Psychiatric Disorders After Traumatic Brain Injury: A 30-Year Follow-Up Study. American Journal of Psychiatry 2002; 159(8): 1315–21.

21. Loignon A, Ouellet M-C, Belleville G. A Systematic Review and Meta-analysis on PTSD Following TBI Among Military/Veteran and Civilian Populations. The Journal of Head Trauma Rehabilitation 2020; 35(1): E21–E35.

22. Dehbozorgi M, Maghsoudi MR, Mohammadi I, et al. Incidence of anxiety after traumatic brain injury: a systematic review and meta-analysis. BMC Neurology 2024; 24(1): 293.

23. Aldossary NM, Kotb MA, Kamal AM. Predictive value of early MRI findings on neurocognitive and psychiatric outcomes in patients with severe traumatic brain injury. Journal of Affective Disorders 2019; 243: 1–7.

24. Shahim P, Politis A, van der Merwe A, et al. Time course and diagnostic utility of NfL, tau, GFAP, and UCH-L1 in subacute and chronic TBI. Neurology 2020; 95(6): e623–e36.

25. Ciryam P, Gerzanich V, Simard JM. Interleukin-6 in Traumatic Brain Injury: A Janus-Faced Player in Damage and Repair. J Neurotrauma 2023; 40(21-22): 2249–69.

26. Korley FK, Jain S, Sun X, et al. Prognostic value of day-of-injury plasma GFAP and UCH-L1 concentrations for predicting functional recovery after traumatic brain injury in patients from the US TRACK-TBI cohort: an observational cohort study. Lancet Neurol 2022; 21(9): 803–13.

27. Li G, Iliff J, Shofer J, et al. CSF β-Amyloid and Tau Biomarker Changes in Veterans With Mild Traumatic Brain Injury. Neurology 2024; 102(7): e209197.

28. Li B, Fang Y, Lin J, Chen X, Li C, He M. Forensic psychiatric analysis of organic personality disorders after craniocerebral injury in Shanghai, China. Frontiers in psychiatry 2022; 13: 944888.

29. Zhang Y, Xiong Y, Yu Q, Shen S, Chen L, Lei X. The activity of daily living (ADL) subgroups and health impairment among Chinese elderly: a latent profile analysis. BMC Geriatrics 2021; 21(1): 30.

30. Hong X, Currier GW, Zhao X, Jiang Y, Zhou W, Wei J. Posttraumatic stress disorder in convalescent severe acute respiratory syndrome patients: a 4-year follow-up study. General Hospital Psychiatry 2009; 31(6): 546–54.

31. Pereira AR, Sanchez-Peña P, Biondi A, et al. Predictors of 1-year outcome after coiling for poor-grade subarachnoid aneurysmal hemorrhage. Neurocritical Care 2007; 7(1): 18–26.

32. Chen YF. Chinese classification of mental disorders (CCMD-3): towards integration in international classification. Psychopathology 2002; 35(2-3): 171–5.

33. Voormolen DC, Cnossen MC, Polinder S, von Steinbuechel N, Vos PE, Haagsma JA. Divergent Classification Methods of Post-Concussion Syndrome after Mild Traumatic Brain Injury: Prevalence Rates, Risk Factors, and Functional Outcome. Journal of Neurotrauma 2018; 35(11): 1233–41.

34. Nelson LD, Kramer MD, Joyner KJ, et al. Relationship between transdiagnostic dimensions of psychopathology and traumatic brain injury (TBI): A TRACK-TBI study. Journal of abnormal psychology 2021; 130(5): 423.

35. Howlett JR, Nelson LD, Stein MB. Mental Health Consequences of Traumatic Brain Injury. Biol Psychiatry 2022; 91(5): 413–20.

36. Sherer M, Struchen MA, Yablon SA, Wang Y, Nick TG. Comparison of indices of traumatic brain injury severity: Glasgow Coma Scale, length of coma and post-traumatic amnesia. *Journal of Neurology*, Neurosurgery & Psychiatry 2008; 79(6): 678–85.

37. Kim JH, Ahn JH, Oh JK, Song JH, Park SW, Chang IB. Factors associated with the development and outcome of hydrocephalus after decompressive craniectomy for traumatic brain injury. Neurosurgical Review 2021; 44(1): 471–8.

38. Xie W, Chapeton JI, Bhasin S, et al. The medial temporal lobe supports the quality of visual short-term memory representation. Nature Human Behaviour 2023; 7(4): 627–41.

39. Tuckute G, Paunov A, Kean H, et al. Frontal language areas do not emerge in the absence of temporal language areas: A case study of an individual born without a left temporal lobe. Neuropsychologia 2022; 169: 108184.

40. Olson IR, Berryhill M. Some surprising findings on the involvement of the parietal lobe in human memory. Neurobiology of learning and memory 2009; 91(2): 155–65.

41. Coslett HB, Schwartz MF. Chapter 18 - The parietal lobe and language. In: Vallar G, Coslett HB, eds. Handbook of Clinical Neurology: Elsevier; 2018: 365–75.

42. Taing AS, Mundy ME, Ponsford JL, Spitz G. Temporal lobe activation during episodic memory encoding following traumatic brain injury. Sci Rep 2021; 11(1): 18830.

43. Dronkers NF. A new brain region for coordinating speech articulation. Nature 1996; 384(6605): 159–61.

44. Heyer DB, Wilbers R, Galakhova AA, et al. Verbal and General IQ Associate with Supragranular Layer Thickness and Cell Properties of the Left Temporal Cortex. Cereb Cortex 2022; 32(11): 2343–57.

45. Rodney T, Taylor P, Dunbar K, et al. High IL-6 in military personnel relates to multiple traumatic brain injuries and post-traumatic stress disorder. Behavioural brain research 2020; 392: 112715.

46. Mendez MF. What is the Relationship of Traumatic Brain Injury to Dementia? Journal of Alzheimer’s disease : JAD 2017; 57(3): 667–81.

47. Rubenstein R, Chang B, Yue JK, et al. Comparing Plasma Phospho Tau, Total Tau, and Phospho Tau-Total Tau Ratio as Acute and Chronic Traumatic Brain Injury Biomarkers. JAMA neurology 2017; 74(9): 1063–72.

48. Wong KR, O’Brien WT, Sun M, et al. Serum Neurofilament Light as a Biomarker of Traumatic Brain Injury in the Presence of Concomitant Peripheral Injury. Biomarker insights 2021; 16: 11772719211053449.

49. Middeldorp J, Hol EM. GFAP in health and disease. Progress in neurobiology 2011; 93(3): 421–43.

50. Clairembault T, Kamphuis W, Leclair-Visonneau L, et al. Enteric GFAP expression and phosphorylation in Parkinson’s disease. Journal of neurochemistry 2014; 130(6): 805–15.

