## Supplementary text and figures for "Chronic Neurological and Psychiatric Outcome in TBI Patients Assessed by A classification System Integrating Symptoms and Radiological Evidence"

**Supplementary text 1**

### QUESTIONNAIRE FOR VARIABLES USED IN MENTAL DISABILITY CLASSIFICATION PREDICTION MODEL

**This questionnaire is used to obtain the features in mental disability classification prediction model. There are three parts in this questionnaire. The first one is for eight binary variables, the second one is for degree of basic activities of daily living (BADL) impairment and degree of social functioning impairment, and the third one is for two ordinal variables.**

**Part 1. The features illustrated in the table below are binary variables with two levels (*Yes* and *No*), *Yes* means the patient fits the description while *No* means the patient doesn't fit this description.**

| Feature | Level | Positive situation (once occurrent the situation below, the level is *Yes*) |
|---|---|---|
| Hypobulia or hyperbulia | *Yes*/*No* | Experience a lack of interest or initiative in pursuing goals, tasks, or activities; or display an intense and overwhelming urge to engage in goal-directed behavior, often to the point of being compulsive or obsessive. |
| Emotional dysregulation | *Yes*/*No* | Inability to regulate the quality and intensity of emotions such as fear, anger, and sadness to produce an appropriate emotional response. |
| Intellectual impairment | *Yes*/*No* | Wechsler Adult Intelligence Scale <=69;<br>In cases of subject unable to complete the above scale, the patient is rated "*Yes*" if he/she is unable to do daily activities independently based on their own intelligence and thinking abilities (excluding cases where hypobulia affects independent daily living). |
| Disorientation | *Yes*/*No* | A state of confusion, uncertainty, or loss of awareness one of one's surroundings, time, or identity. |
| Remote memory impairment (remote memory encompasses long-term memories that were formed months, or even years ago) | *Yes*/*No* | Difficulties in recalling or retrieving memories of events, facts, or experiences that occurred in the distant past. |
| Communication disturbances | *Yes*/*No* | Aphasia or damage to the vocal organs resulting in difficulty in speaking. |
| Nonspecific psychiatric symptom | *Yes*/*No* | One or more symptoms including headache, dizziness, fatigue or weakness, sleep disorders, self-reported memory impairment without clinical evidence, irritability, temper change. |
| Unconsciousness | *Yes*/*No* | A state in which an individual lacks awareness of themselves and their surroundings, as well as the inability to respond to external stimuli. |

**Part 2. Each of the two features illustrated below should be calculated as an overall rated level after individual activity scores are collected. Each activity score corresponds to the level of impairment of a given activity. For each activity score, there are four possible levels (*0, 1, 2* or *unknow*): *0* means the patient doesn't have impairment of this description, *1* means the patient has a partial impairment of the description, *2* means the patient has complete impairment of this description, *unknow* means missing information for this description.**

<table>
<tr><td>Feature</td><td>Degree of BADL impairment (0,1,2)</td><td>Activity score (0,1,2 or unknow)</td><td>Activity</td></tr>
<tr><td rowspan="6">Degree of BADL impairment</td><td rowspan="6"></td><td></td><td>Walking. Patient walks steadily in flat ground.</td></tr>
<tr><td></td><td>Feeding. Patient gets food from plate into mouth without help. Preparation of food may be done by another person.</td></tr>
<tr><td></td><td>Dressing. Patient gets clothes from closets and drawers and puts on clothes and outer garments complete with fasteners.</td></tr>
<tr><td></td><td>Bathing, brushing teeth, washing face. Patient can bath self, brush teeth and wash face.</td></tr>
<tr><td></td><td>Toileting. Patient goes to toilet, gets on and off, arranges clothes, cleans genital area.</td></tr>
<tr><td colspan="2">Once activity scores are collected, only using the activity scores that are not unknown (activity score = 0/1/2) of the activities listed above to rate the overall BADL impairment level as below:<br>Rate 0 when all activity scores are 0.<br>Rate 1: when any of the following situations is satisfied:<br>1) When both 0 and non-0 activity scores are present, and less than 2/3 activities have an activity score of 2<br>2) when only non-0 activity scores are present, and less than 1/3 activities have an activity score of 2<br>Rate 2: when any of the following situations is satisfied:<br>1) When both 0 and non-0 activity scores are present, and greater than or equal to 2/3 activities have an activity score of 2.<br>2) when only non-0 activity scores are present, and greater than or equal to 1/3 activities have an activity score of 2.</td></tr>
</table>

<table>
<tr><th>Feature</th><th>Degree of social functioning impairment (0,1,2)</th><th>Activity score (0,1,2 or unknow)</th><th>Activity</th></tr>
<tr><td rowspan="10">Degree of social functioning impairment</td><td rowspan="10"></td><td></td><td>Work/school tasks</td></tr>
<tr><td></td><td>The ability to get along with spouse</td></tr>
<tr><td></td><td>The ability to get along with children</td></tr>
<tr><td></td><td>The ability to get along with friends and strangers.</td></tr>
<tr><td></td><td>Joining in community activities (for example, shopping, festivities, religious, or other activities)</td></tr>
<tr><td></td><td>Activities inside home</td></tr>
<tr><td></td><td>Household tasks and responsibilities</td></tr>
<tr><td></td><td>Interest and concern for the outside world</td></tr>
<tr><td></td><td>Responsibility and planning</td></tr>
<tr><td colspan="2">Once activity scores are collected, only using the activity scores that are not unknown (activity score = 0/1/2) of the activities listed above to rate the overall social function impairment level as below:<br>Rate 0 when all activity scores are 0.<br>Rate 1: when any of the following situations is satisfied:<br>1) When both 0 and non-0 activity scores are present, and less than 2/3 activities have an activity score of 2<br>2) when only non-0 activity scores are present, and less than 1/3 activities have an activity score of 2<br>Rate 2: when any of the following situations is satisfied:<br>1) When both 0 and non-0 activity scores are present, and greater than or equal to 2/3 activities have an activity score of 2.<br>2) when only non-0 activity scores are present, and greater than or equal to 1/3 activities have an activity score of 2.</td></tr>
</table>

**Part 3. The two features illustrated below are assessed levels. The standard is given in descriptions below.**

| Feature | Level | Description |
|---|---|---|
| The number of lobes with encephalomalacia | | This feature includes four lobes: frontal lobe, temporal lobe, parietal lobe, and occipital lobe, without distinguishing between left and right brain.<br>**Level *0*:** no encephalomalacia<br>**Level *1*:** there is one lobe with encephalomalacia<br>**Level *2*:** there are two lobes with encephalomalacia<br>**Level *3*:** there are three lobes with encephalomalacia<br>**Level *4*:** there are four lobes with encephalomalacia |
| Degree of insight impairment | | It is suggested that insight has at least three dimensions: (a) awareness of illness, (b) the capacity to relabel psychotic experiences as abnormal, and (c)treatment compliance.<br>**Level *0*:** no insight impairment<br>**Level *1*:** partial impairment of insight<br>**Level *2*:** complete impairment of insight |

### Supplementary Figures

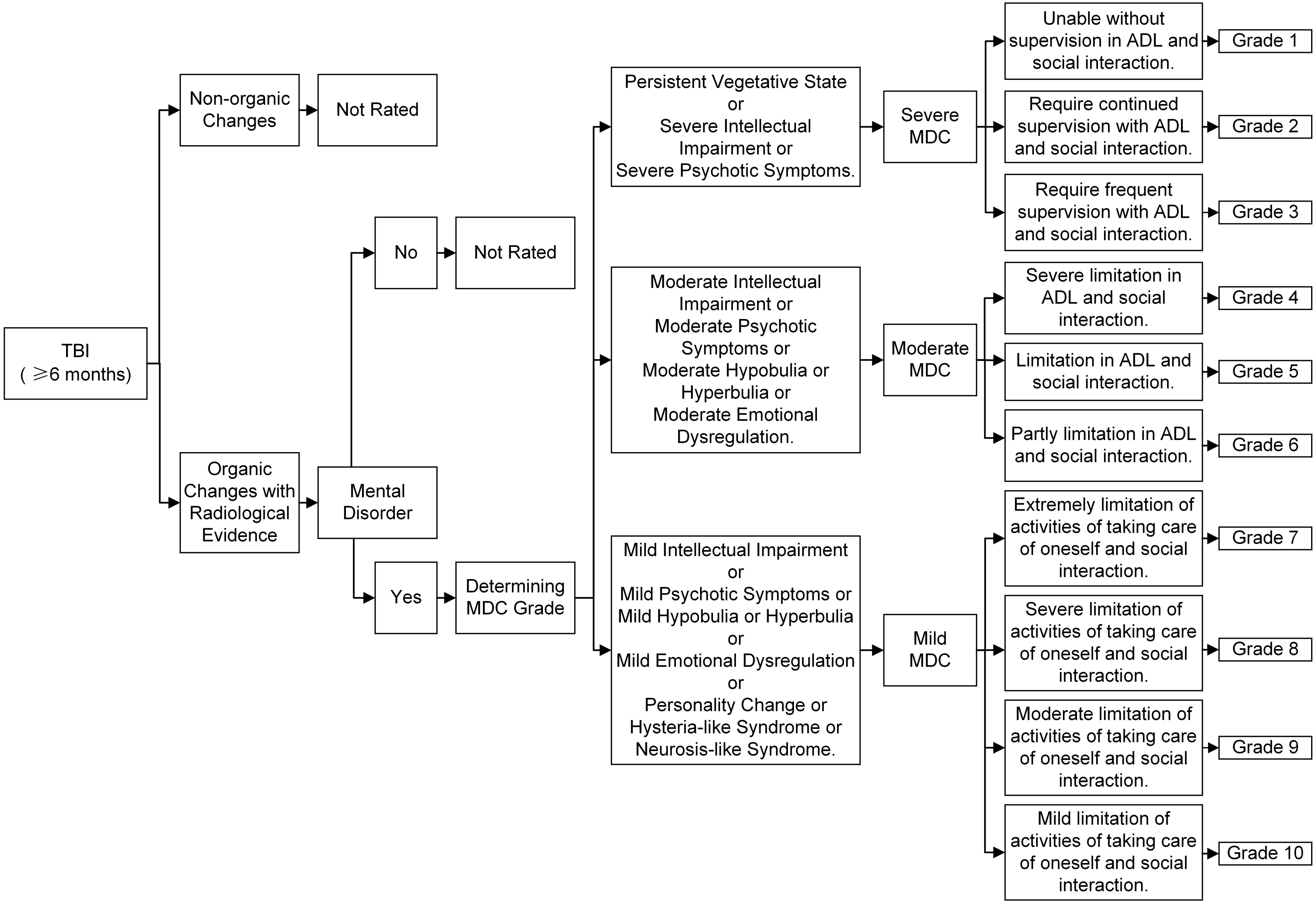


**Supplementary Figure 1 Mental Disability Evaluation Criteria from Guidelines for Mental Disability Classification (MDC) Grade in China**

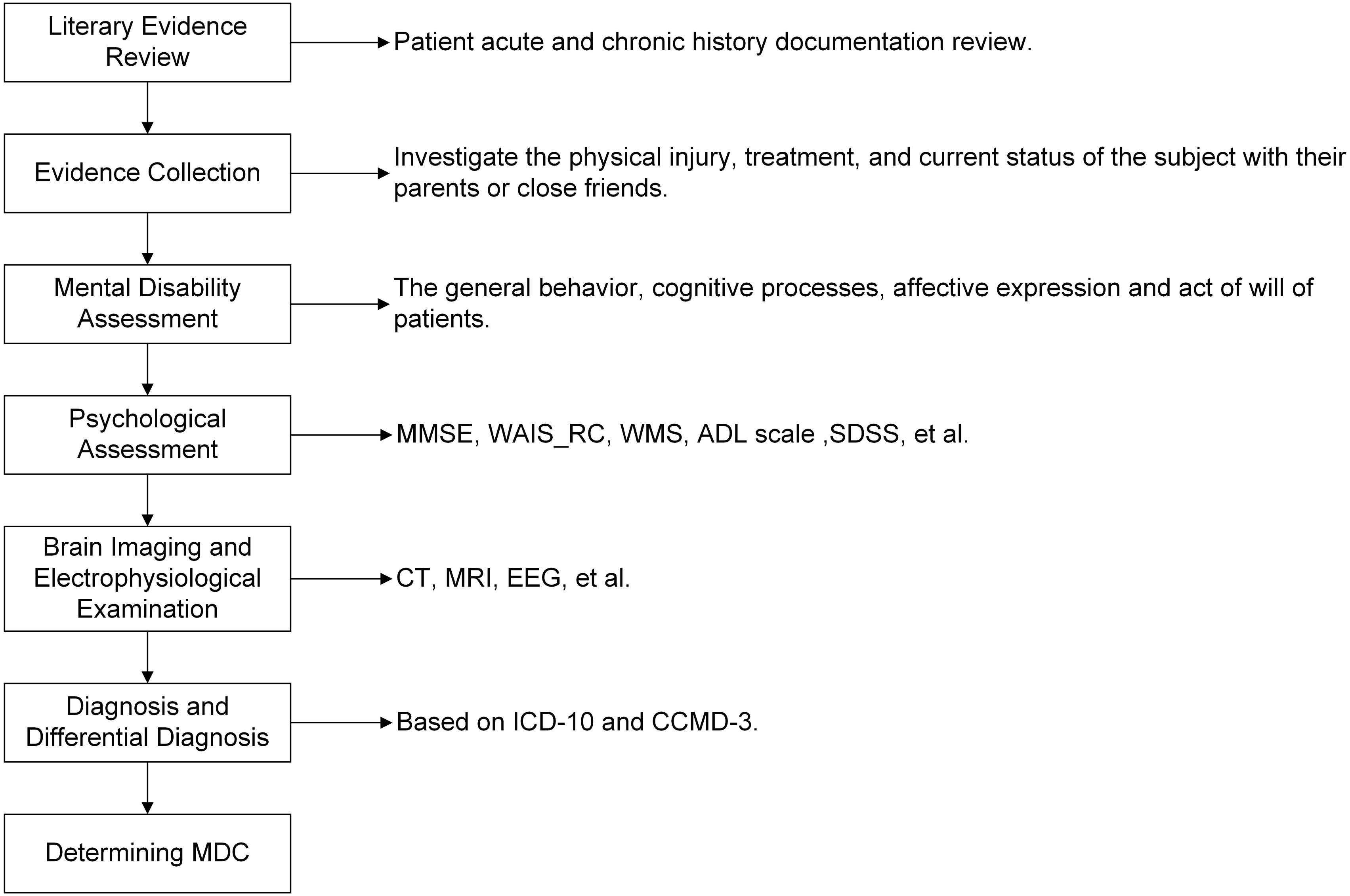


**Supplementary Figure 2 Mental Disability Evaluation Procedure.**

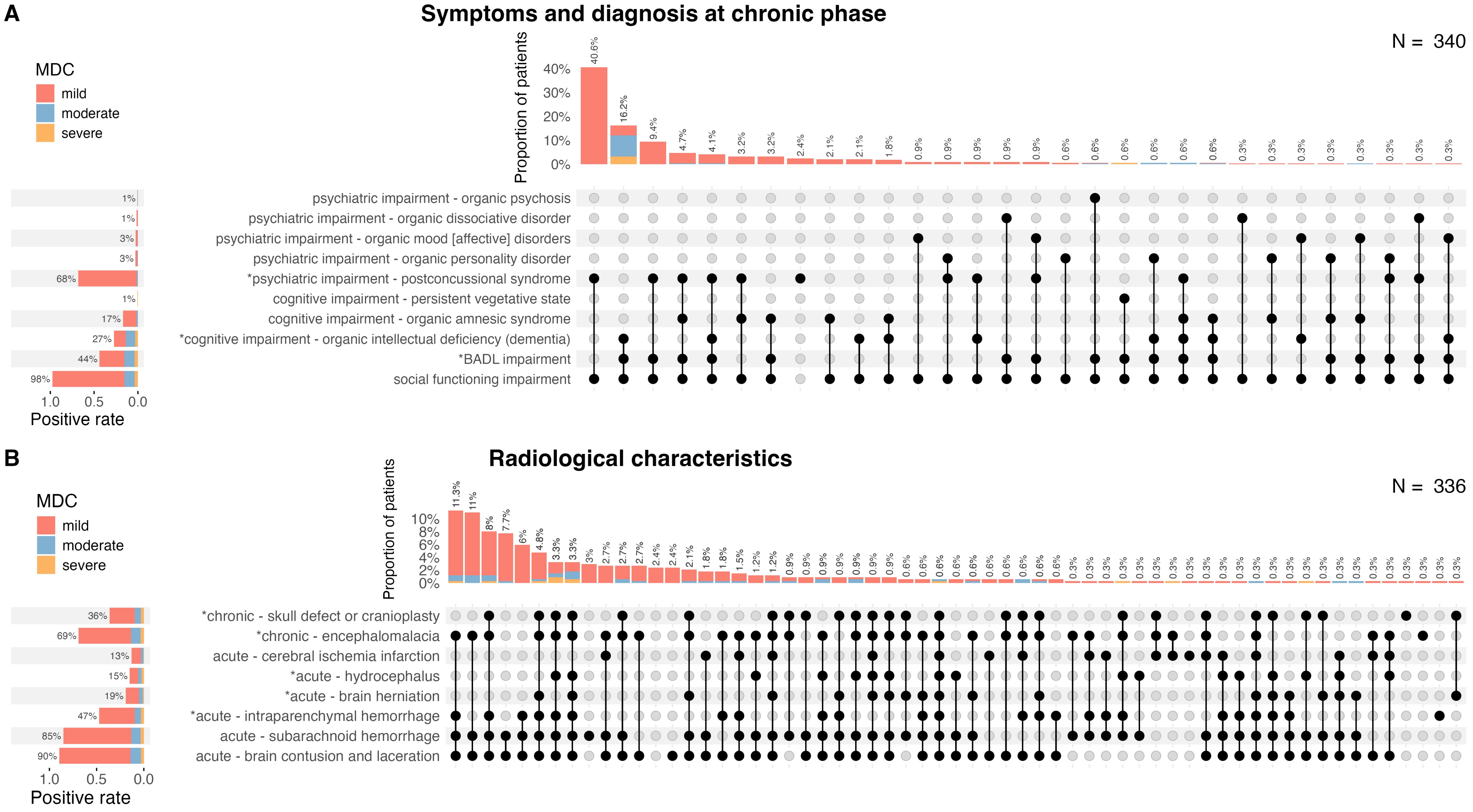


**Supplementary Figure 3 The distribution of different symptoms and diagnosis at chronic phase (A) and radiological characteristics (B) in TBI patients with varied MDC severity.** *: $P_{FDR}$ <0.05

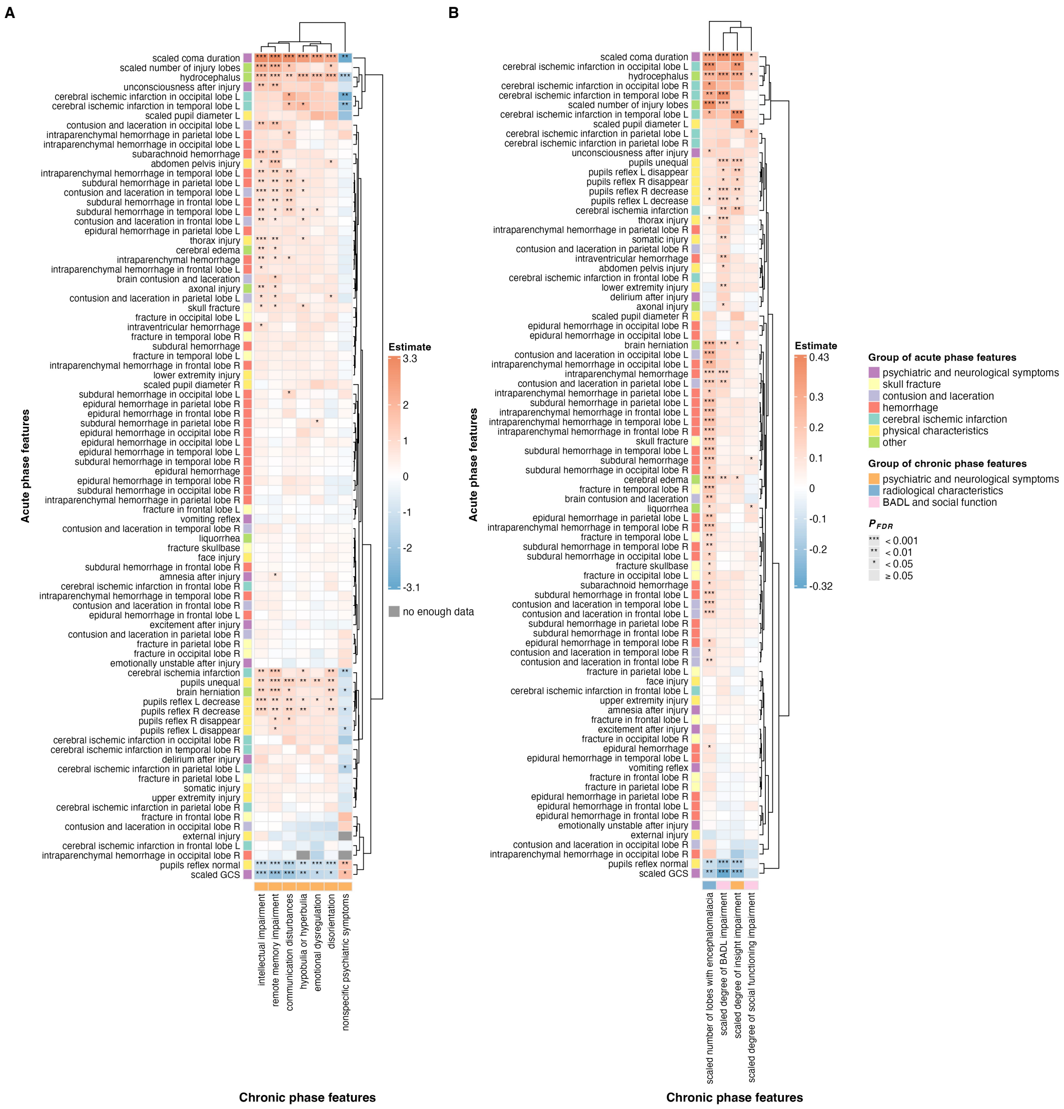


**Supplementary Figure 4 The association results between 86 acute features and 11 deconstructed chronic dimensional features from MDC. A.** The association between 86 acute features and 7 binary chronic features. **B.** The association between 86 acute features and 4 ordinal chronic features.

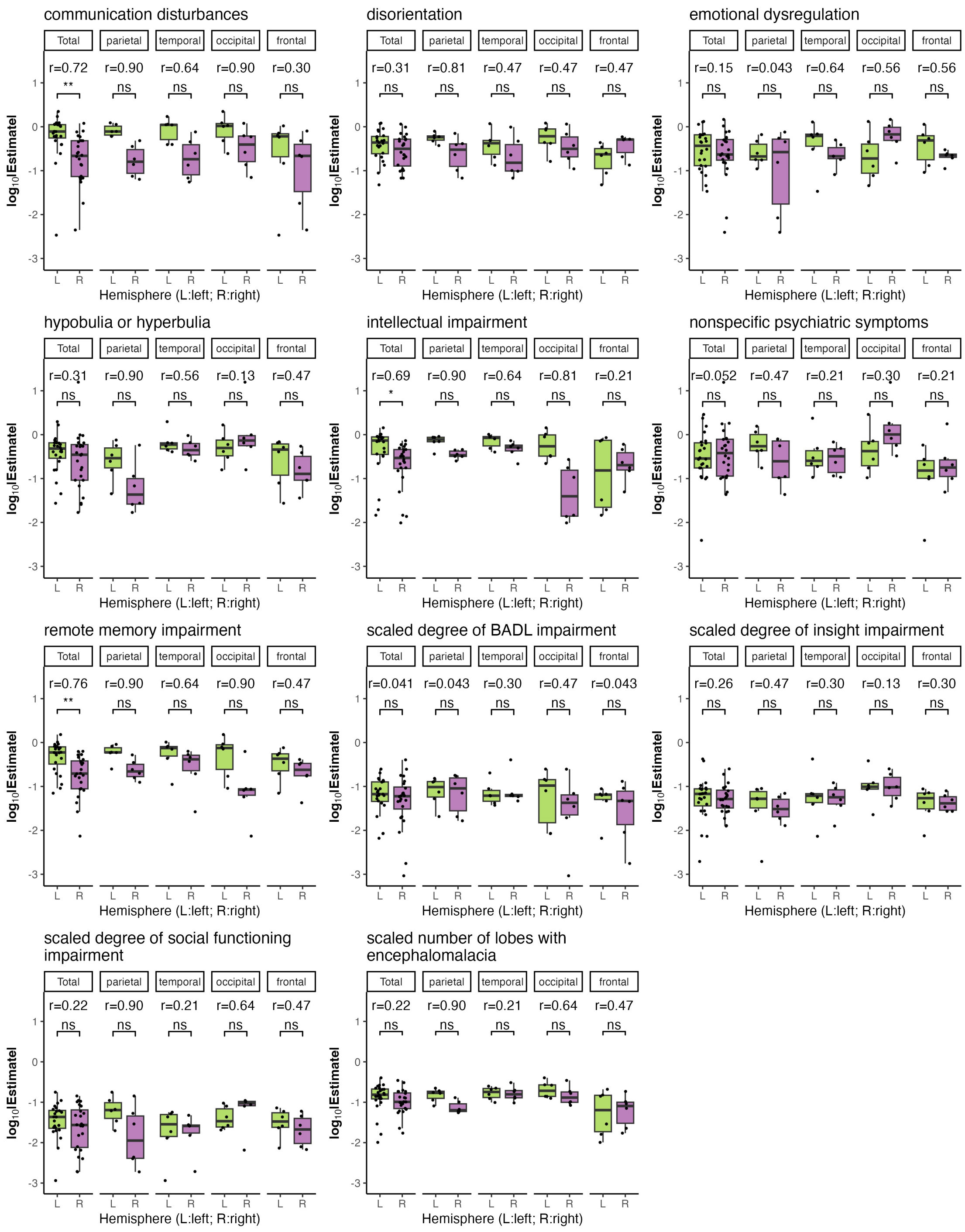


**Supplementary Figure 5 Results of Wilcoxon tests comparing the absolute value of regression coefficient between acute injury in left and right hemispheres for each of the key chronic dimensional features.** r: effect size, $\log_{10}$|Estimate|: the base-10 logarithm of the absolute regression coefficient. ***: $P_{FDR}$<0.0001, **: $P_{FDR}$<0.001, *: $P_{FDR}$<0.05, ns: $P_{FDR}$≥0.05.

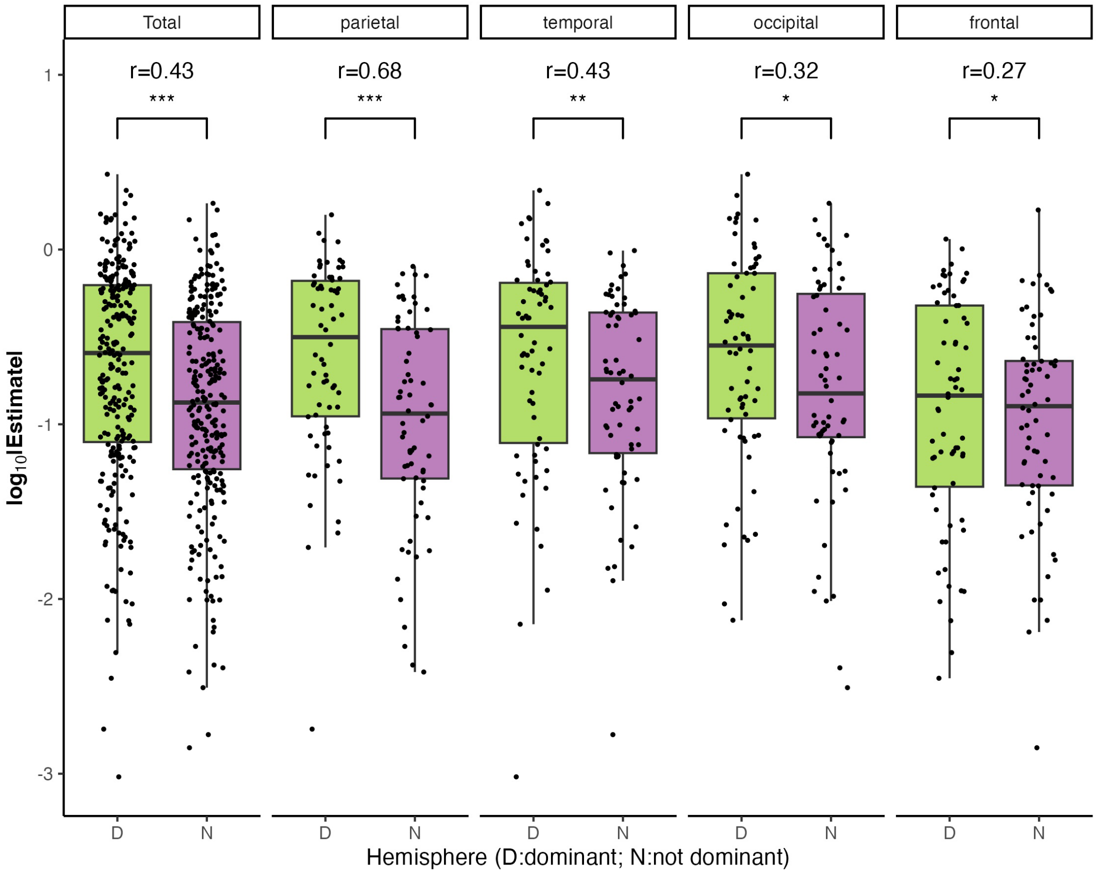


**Supplementary Figure 6** Results of Wilcoxon tests comparing the absolute value of regression coefficient between chronic associations of acute injuries in dominant and non-dominant hemispheres. r: effect size, $\log_{10}$|Estimate|: the base-10 logarithm of the absolute regression coefficient. ***: $P_{FDR}$<0.0001, **: $P_{FDR}$<0.001, *: $P_{FDR}$<0.05, ns: $P_{FDR}\geq$0.05.

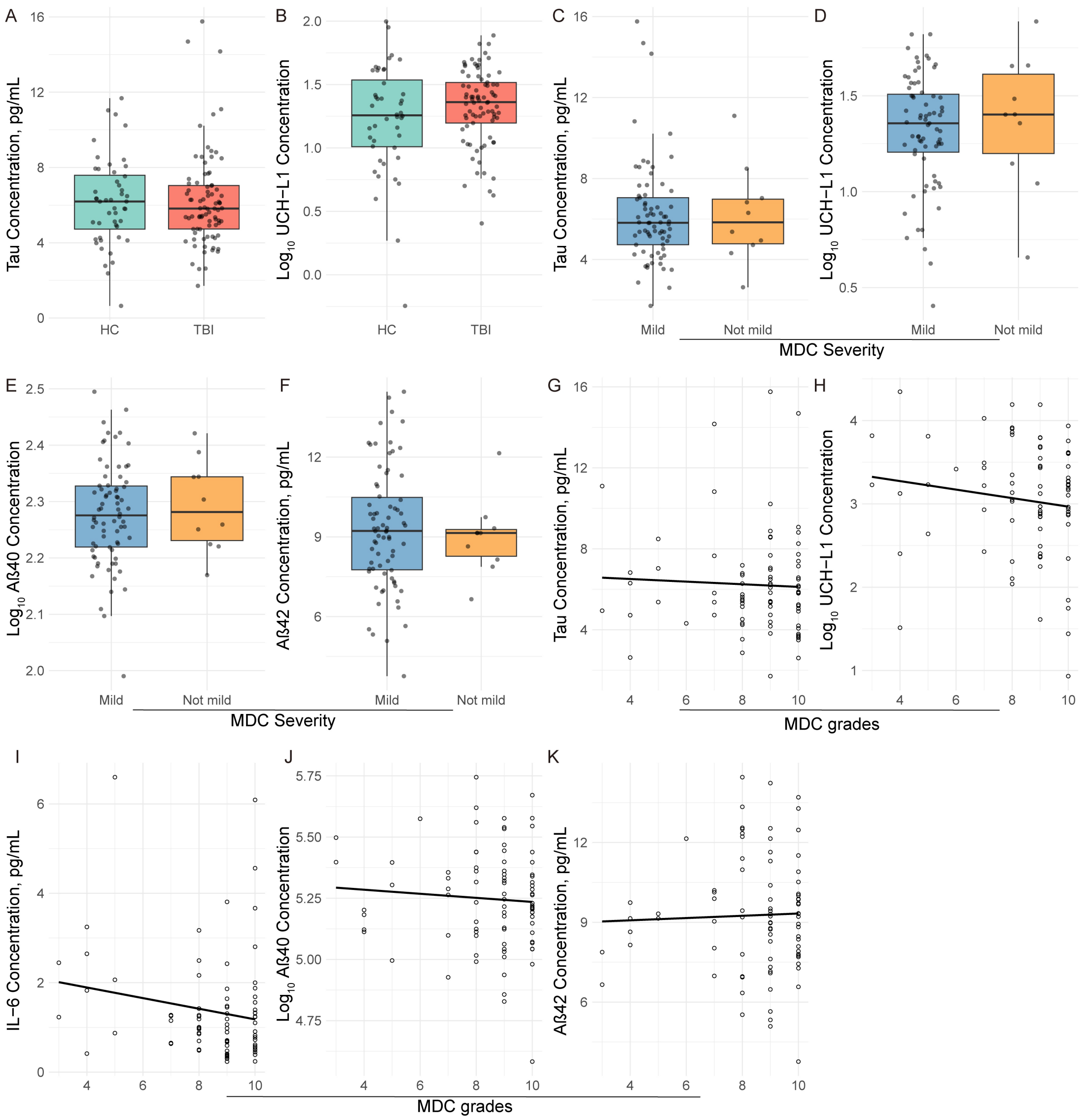


**Supplementary Figure 7 Non-significant results from associations tests between protein concentration and mental prognosis of TBI.** A and B: Comparisons of protein concentration between TBI and HC groups; C to F: Comparisons of protein concentration across MDC severity groups; G to K: Association between protein concentration and MDC grades.

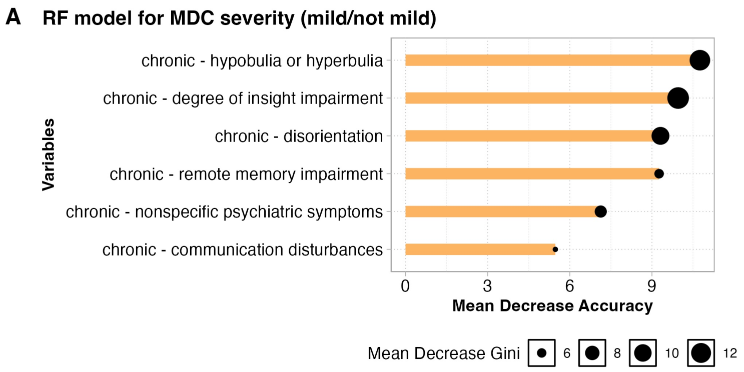


| Cohorts | Ncases | Algorithm | AUC | Sensitivity | Specificity | Accuracy |
|---|---|---|---|---|---|---|
| SYSU-TBI1 | 356 | RF | 0.9751 | 0.9138 | 0.9362 | 0.9326 |
| SYSU-TBI2 | 84 | RF | 0.9730 | 0.8000 | 0.9459 | 0.9286 |

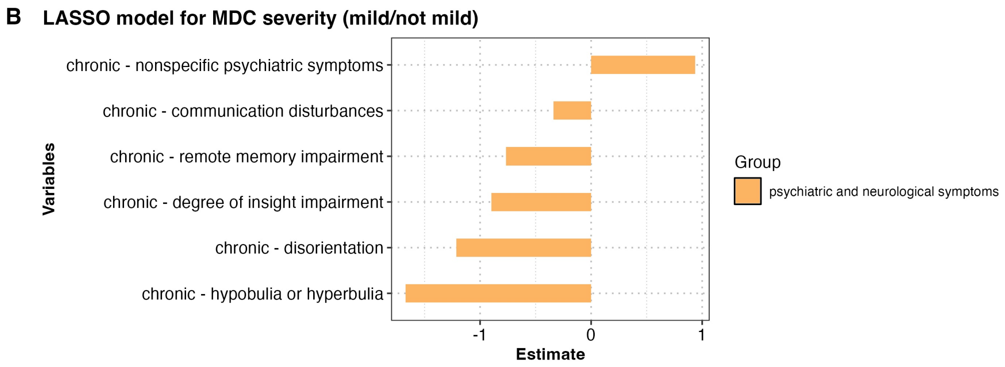


| Cohorts | Ncases | Algorithm | AUC | Sensitivity | Specificity | Accuracy |
|---|---|---|---|---|---|---|
| SYSU-TBI1 | 356 | LASSO | 0.9796 | 0.9732 | 0.7586 | 0.9382 |
| SYSU-TBI2 | 84 | LASSO | 0.9784 | 0.9730 | 0.6000 | 0.9286 |

**Supplementary Figure 8 The variables of prediction models for MDC severity (mild/not mild). A.** The variables importance of the RF prediction model for MDC severity (mild/not mild). **B.** The variables regression coefficient of the LASSO prediction model for MDC severity (mild/not mild).

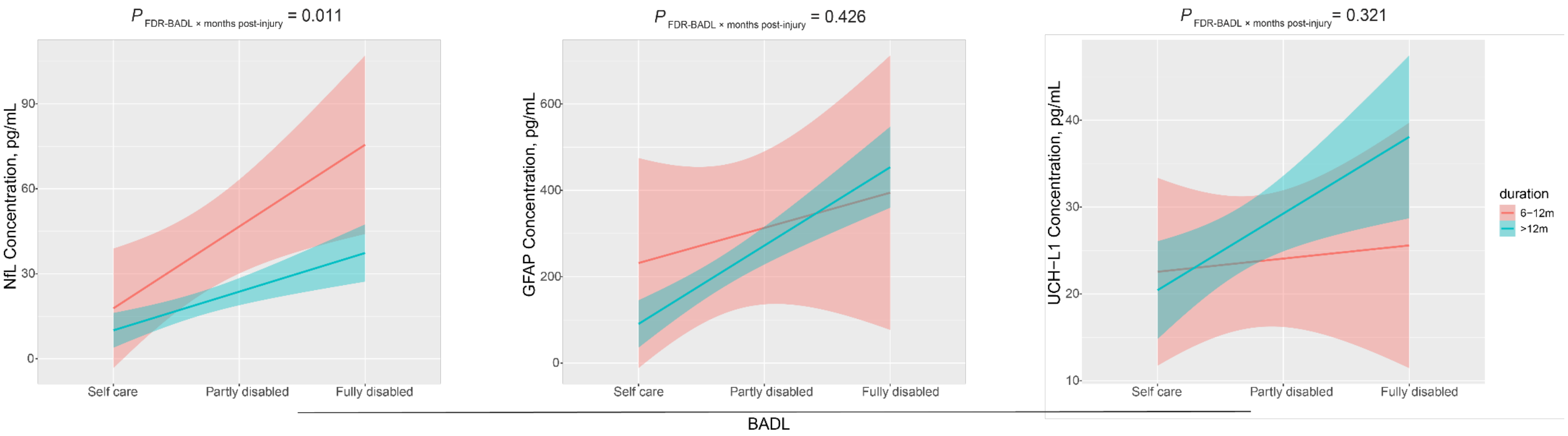


**Supplementary Figure 9** Interactive effect between BADL and months post-injury on NfL, GFAP and UCH-L1**.** Months post-injury was treated as a continuous variable in interactive effect analyses.
